# Meta-analysis identifies the patient classification major impact on the ACPA association with rheumatoid arthritis-associated interstitial lung disease (RA-ILD)

**DOI:** 10.1101/2025.02.24.25322770

**Authors:** Bartosz Kaczmarczyk, Carmen Conde, Antonio Gonzalez, MARILD network

## Abstract

**Background:** We need screening for RA patients at high risk of RA-ILD to prevent the associated decrease in life quality and survival. The proposed screenings disagree regarding the anti-citrullinated protein antibodies (ACPA) because of their inconsistent association across recent studies. Therefore, we hypothesized that meta-analysis of the published reports should reveal clues explaining the heterogeneity of results and that knowing them could help us progress in RA-ILD early detection.

**Objectives:** We aimed to discover the factors accounting for the variability of the ACPA association in the published reports.

**Methods:** We searched the Web of Science and PubMed databases for studies reporting ACPA in RA-ILD and RA-control groups. The identified studies were analyzed using meta-analysis and meta-regression to identify moderators of the ACPA association.

**Results:** We found 513 unique records, containing 31 eligible data sets. The meta-analysis preceding the search for moderators showed a remarkable heterogeneity (p_Q_ = 5.7 x 10^-7^). Appropriate tests showed that it was largely attributable (58.1 %) to an outlier study, which had recruited cases and controls in different place and time contexts. The exclusion of this outlier from subsequent analyses did not completely remove heterogeneity (p_Q_ = 0.004). However, it permitted the identification of the patient classification method as a significant moderator: The 14 studies using chest CT showed stronger ACPA association with RA-ILD (OR = 3.05 [95%CI: 2.12-4.38]) than the 16 employing multifactorial criteria (1.55 [95%CI: 1.18-2.03]; p = 0.0047 for the contrast). This moderator accounted for the significant heterogeneity (p_Q_ = 0.079), was robust in sensitivity analyses, and was the only one found.

**Conclusions:** Our results validate the ACPA association with RA-ILD, reinforce the importance of study design, and suggest the need to consider if studies relying on chest CT for classification could be more fruitful in the search for RA-ILD biomarkers.

## Background

Rheumatoid arthritis-associated interstitial lung disease (RA-ILD) appears as a clinically significant complication in about 10 % of the patients [1, 2]. It courses with dry cough and dyspnea associated with lung fibrosis, impaired pulmonary functionality, a decrease in life quality, and premature death. Fortunately, the perspectives of the affected patients are improving thanks to the growing awareness and the new anti-fibrotic drugs. Current recommendations demand the evaluation of all RA patients for ILD risk, to follow the high-risk patients with increased periodicity and attention [2-5]. Additional progress requires the development of screening instruments to estimate RA-ILD risk. We already know a set of risk factors that include old age, male sex, smoking, the risk allele of a *MUC5B* polymorphism, and the presence of anti-citrullinated protein antibodies (ACPA). However, the latter has been questioned because some large studies did not observe ACPA association with RA-ILD [3, 6]. The role of ACPAs requires clarification because they are a component of some proposed RA-ILD screening tools but not of others [3-5, 7, 8].

Regarding the discordant results, there is a suspicion that risk factors could vary depending on multiple factors, like the RA-ILD classification or stage, the use of ACPA to select RA patients, and the design of the study [3, 9, 10]. Some of these factors could be relevant for other RA-ILD biomarkers, increasing the interest in discovering their impact on the ACPA association with RA-ILD [1, 11].

The previously published meta-analyses do not solve the doubts about ACPA because they were not focused on the search for the causes of the studies’ heterogeneity [12-16]. All meta-analyses showed a significant association of ACPA with RA-ILD. However, the summary effect sizes they reported show a striking decline with time: from OR = 4.68 in the meta-analysis published in 2014 to OR = 1.58 in 2023 [15, 16]. This decline has been unadverted and has not been investigated. Therefore, we decided to focus on the causes of the heterogeneity and decline with time using meta-analysis and meta-regression methods. The utility of meta-analysis with this objective is well-established [17-21], although, less common than summarizing the available studies. To facilitate the understanding, we have added some explanations that we expect could compensate for the lower familiarity with this application of meta-analysis.

## Methods

### Bibliographic search strategy and study selection

The literature review was conducted according to the Preferred Reporting items for systematic Reviews and Meta-analyses guidelines [22]. The PubMed and Web of Science databases were queried for publications containing “(rheumatoid arthritis-associated interstitial lung disease OR ra-ild) AND (acpa OR anti-cpp OR anti-citrullinated protein antibodies OR anti-citrullinated peptide antibodies OR risk factors)” and published in the period from the 1st January 2000 to the 1st May 2024. In addition, the references of the selected reports and the previously published meta-analyses were also explored searching for any study that could have been missed. Article selection, classification, and data extraction are described in the Suppl. Methods.

### Meta-analysis and meta-regression to identify moderators

We used random effect meta-analysis with inverse variance weights and maximum likelihood estimation [17-21]. Maximum likelihood was selected because it allows tests for model selection [17, 19, 20]. Assessment of the meta-analysis included a search for outliers, small study bias, and baseline effects by using, respectively, the influence measures included in the *metafor* package [20]; the funnel plot and statistical tests; and the baseline plot. The qualitative variables were treated as dichotomous; when significant subgroup meta-analysis was also performed. The quantitative variables showing a skewed distribution were transformed. The software used were the *metafor-based* JASP (v0.18.3) and R (v4.3.2). P-values < 0.05 were considered statistically significant. Additional information on the analysis methods is provided in the Suppl. Methods.

## Results

### The bibliography search identified 31 eligible data sets

The bibliography search identified 513 non-redundant records containing the query terms (Fig. 1). The reviews, case reports, records not in English, retracted publications, and patents were excluded. The remaining 381 records were retrieved and independently assessed by two authors. A large fraction of them did not contain the required information, mainly because their design did not include RA-ILD and RA-control groups. Instead, they reported time-to-event analyses, comparisons of subgroups within RA-ILD, or contrasts between RA-ILD and other diseases or healthy controls. Additionally, some records reported frequencies of RF-positive or seropositive patients without specifying the ACPA frequency. Therefore, only 30 records containing 31 data sets were selected for meta-analysis (Table 1 and Suppl. Table 1). One was a meeting abstract [23], and the remaining were journal articles (Suppl. References). All were published between 2012 and 2023.

**Figure 1:**
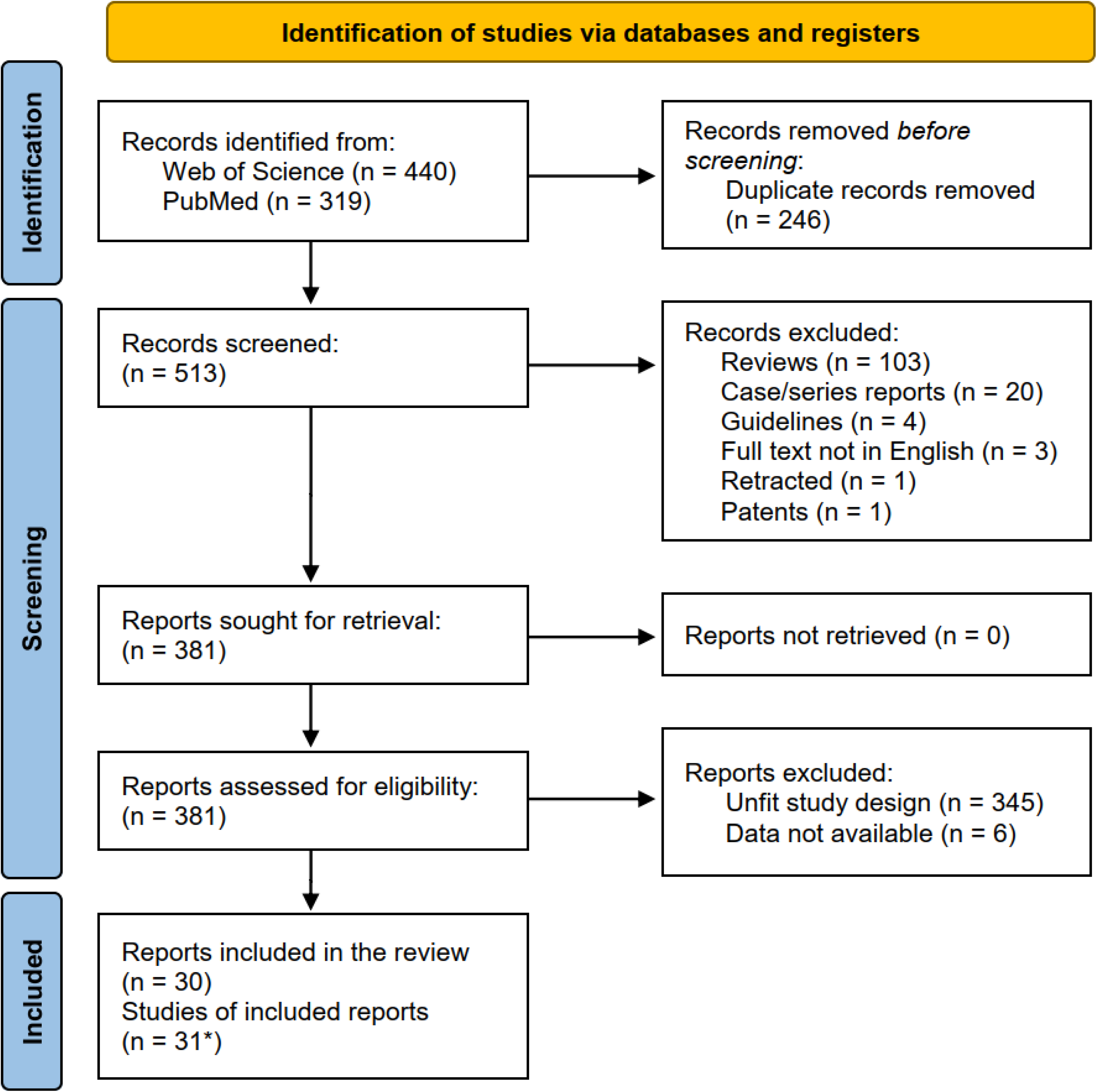
Flow diagram of identification, filtering, and selection of the reports.

**Table 1.**
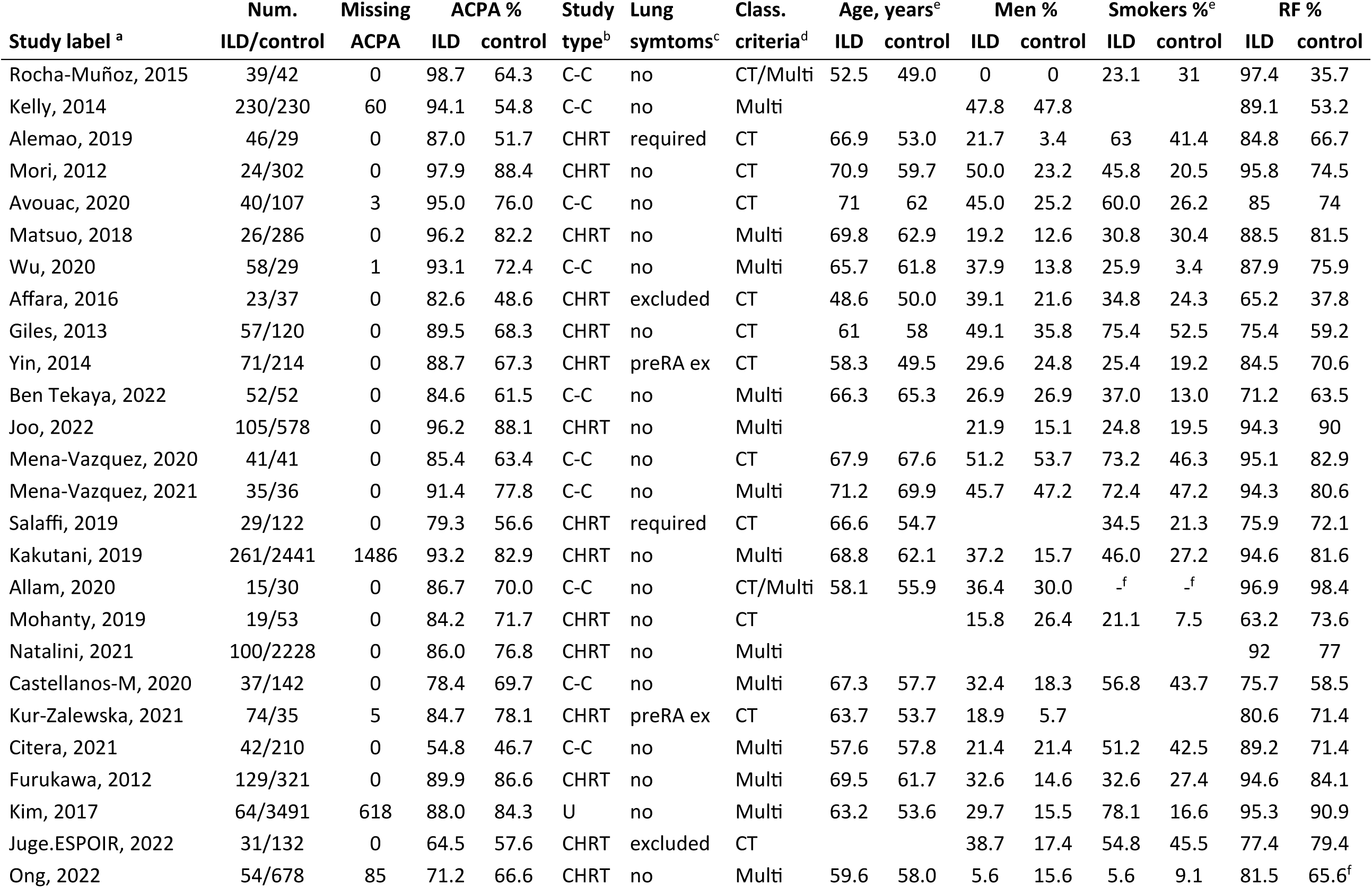
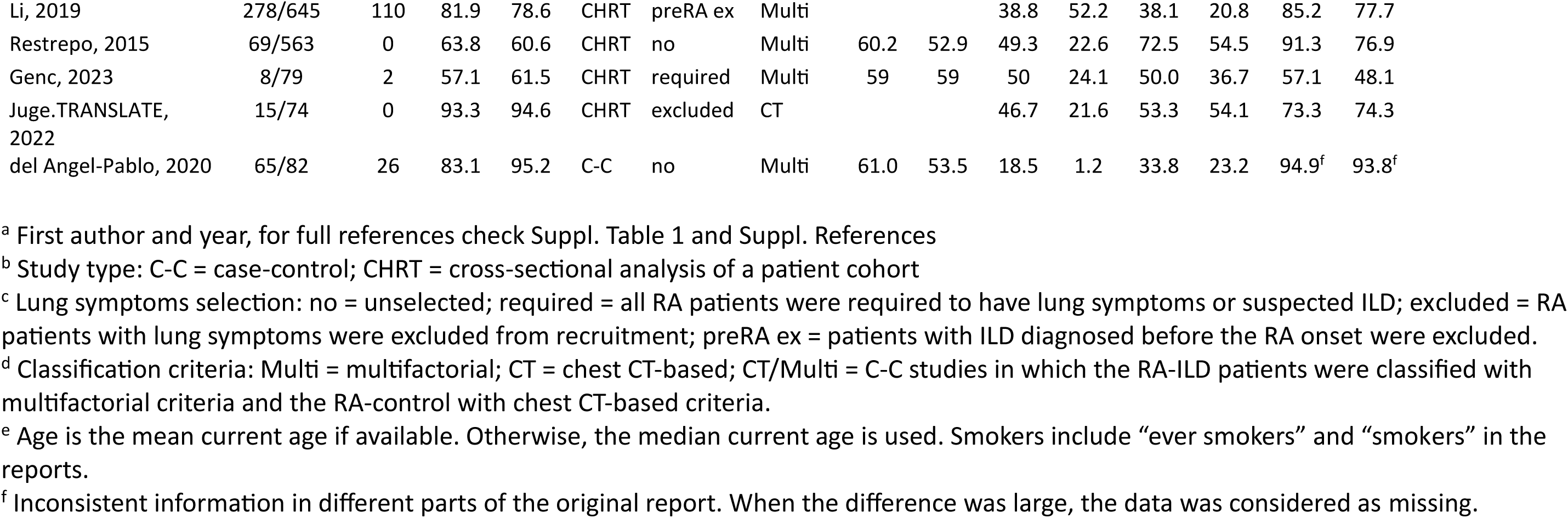
Main characteristics of the 31 studies selected for meta-analysis.

| Study label <sup>a</sup> | Num. | Missing | ACPA % |  | Study | Lung | Class. | Age, years<sup>e</sup> |  | Men % |  | Smokers %<sup>e</sup> |  | RF % |  |
| --- | --- | --- | --- | --- | --- | --- | --- | --- | --- | --- | --- | --- | --- | --- | --- |
|  | ILD/control | ACPA | ILD | control | type<sup>b</sup> | syntoms<sup>c</sup> | criteria<sup>d</sup> | ILD | control | ILD | control | ILD | control | ILD | control |
| Rocha-Muñoz, 2015 | 39/42 | 0 | 98.7 | 64.3 | C-C | no | CT/Multi | 52.5 | 49.0 | 0 | 0 | 23.1 | 31 | 97.4 | 35.7 |
| Kelly, 2014 | 230/230 | 60 | 94.1 | 54.8 | C-C | no | Multi |  |  | 47.8 | 47.8 |  |  | 89.1 | 53.2 |
| Alemao, 2019 | 46/29 | 0 | 87.0 | 51.7 | CHRT | required | CT | 66.9 | 53.0 | 21.7 | 3.4 | 63 | 41.4 | 84.8 | 66.7 |
| Mori, 2012 | 24/302 | 0 | 97.9 | 88.4 | CHRT | no | CT | 70.9 | 59.7 | 50.0 | 23.2 | 45.8 | 20.5 | 95.8 | 74.5 |
| Avouac, 2020 | 40/107 | 3 | 95.0 | 76.0 | C-C | no | CT | 71 | 62 | 45.0 | 25.2 | 60.0 | 26.2 | 85 | 74 |
| Matsuo, 2018 | 26/286 | 0 | 96.2 | 82.2 | CHRT | no | Multi | 69.8 | 62.9 | 19.2 | 12.6 | 30.8 | 30.4 | 88.5 | 81.5 |
| Wu, 2020 | 58/29 | 1 | 93.1 | 72.4 | C-C | no | Multi | 65.7 | 61.8 | 37.9 | 13.8 | 25.9 | 3.4 | 87.9 | 75.9 |
| Affara, 2016 | 23/37 | 0 | 82.6 | 48.6 | CHRT | excluded | CT | 48.6 | 50.0 | 39.1 | 21.6 | 34.8 | 24.3 | 65.2 | 37.8 |
| Giles, 2013 | 57/120 | 0 | 89.5 | 68.3 | CHRT | no | CT | 61 | 58 | 49.1 | 35.8 | 75.4 | 52.5 | 75.4 | 59.2 |
| Yin, 2014 | 71/214 | 0 | 88.7 | 67.3 | CHRT | preRA ex | CT | 58.3 | 49.5 | 29.6 | 24.8 | 25.4 | 19.2 | 84.5 | 70.6 |
| Ben Tekaya, 2022 | 52/52 | 0 | 84.6 | 61.5 | C-C | no | Multi | 66.3 | 65.3 | 26.9 | 26.9 | 37.0 | 13.0 | 71.2 | 63.5 |
| Joo, 2022 | 105/578 | 0 | 96.2 | 88.1 | CHRT | no | Multi |  |  | 21.9 | 15.1 | 24.8 | 19.5 | 94.3 | 90 |
| Mena-Vazquez, 2020 | 41/41 | 0 | 85.4 | 63.4 | C-C | no | CT | 67.9 | 67.6 | 51.2 | 53.7 | 73.2 | 46.3 | 95.1 | 82.9 |
| Mena-Vazquez, 2021 | 35/36 | 0 | 91.4 | 77.8 | C-C | no | Multi | 71.2 | 69.9 | 45.7 | 47.2 | 72.4 | 47.2 | 94.3 | 80.6 |
| Salaffi, 2019 | 29/122 | 0 | 79.3 | 56.6 | CHRT | required | CT | 66.6 | 54.7 |  |  | 34.5 | 21.3 | 75.9 | 72.1 |
| Kakutani, 2019 | 261/2441 | 1486 | 93.2 | 82.9 | CHRT | no | Multi | 68.8 | 62.1 | 37.2 | 15.7 | 46.0 | 27.2 | 94.6 | 81.6 |
| Allam, 2020 | 15/30 | 0 | 86.7 | 70.0 | C-C | no | CT/Multi | 58.1 | 55.9 | 36.4 | 30.0 | -<sup>f</sup> | -<sup>f</sup> | 96.9 | 98.4 |
| Mohanty, 2019 | 19/53 | 0 | 84.2 | 71.7 | CHRT | no | CT |  |  | 15.8 | 26.4 | 21.1 | 7.5 | 63.2 | 73.6 |
| Natalini, 2021 | 100/2228 | 0 | 86.0 | 76.8 | CHRT | no | Multi |  |  |  |  |  |  | 92 | 77 |
| Castellanos-M, 2020 | 37/142 | 0 | 78.4 | 69.7 | C-C | no | Multi | 67.3 | 57.7 | 32.4 | 18.3 | 56.8 | 43.7 | 75.7 | 58.5 |
| Kur-Zalewska, 2021 | 74/35 | 5 | 84.7 | 78.1 | CHRT | preRA ex | CT | 63.7 | 53.7 | 18.9 | 5.7 |  |  | 80.6 | 71.4 |
| Citera, 2021 | 42/210 | 0 | 54.8 | 46.7 | C-C | no | Multi | 57.6 | 57.8 | 21.4 | 21.4 | 51.2 | 42.5 | 89.2 | 71.4 |
| Furukawa, 2012 | 129/321 | 0 | 89.9 | 86.6 | CHRT | no | Multi | 69.5 | 61.7 | 32.6 | 14.6 | 32.6 | 27.4 | 94.6 | 84.1 |
| Kim, 2017 | 64/3491 | 618 | 88.0 | 84.3 | U | no | Multi | 63.2 | 53.6 | 29.7 | 15.5 | 78.1 | 16.6 | 95.3 | 90.9 |
| Juge.ESPOIR, 2022 | 31/132 | 0 | 64.5 | 57.6 | CHRT | excluded | CT |  |  | 38.7 | 17.4 | 54.8 | 45.5 | 77.4 | 79.4 |
| Ong, 2022 | 54/678 | 85 | 71.2 | 66.6 | CHRT | no | Multi | 59.6 | 58.0 | 5.6 | 15.6 | 5.6 | 9.1 | 81.5 | 65.6<sup>f</sup> |
| Li, 2019 | 278/645 | 110 | 81.9 | 78.6 | CHRT | preRA ex | Multi |  |  | 38.8 | 52.2 | 38.1 | 20.8 | 85.2 | 77.7 |
| Restrepo, 2015 | 69/563 | 0 | 63.8 | 60.6 | CHRT | no | Multi | 60.2 | 52.9 | 49.3 | 22.6 | 72.5 | 54.5 | 91.3 | 76.9 |
| Genc, 2023 | 8/79 | 2 | 57.1 | 61.5 | CHRT | required | Multi | 59 | 59 | 50 | 24.1 | 50.0 | 36.7 | 57.1 | 48.1 |
| Juge.TRANSLATE, 2022 | 15/74 | 0 | 93.3 | 94.6 | CHRT | excluded | CT |  |  | 46.7 | 21.6 | 53.3 | 54.1 | 73.3 | 74.3 |
| del Angel-Pablo, 2020 | 65/82 | 26 | 83.1 | 95.2 | C-C | no | Multi | 61.0 | 53.5 | 18.5 | 1.2 | 33.8 | 23.2 | 94.9<sup>f</sup> | 93.8<sup>f</sup> |

<sup>a</sup> First author and year, for full references check Suppl. Table 1 and Suppl. References
<sup>b</sup> Study type: C-C = case-control; CHRT = cross-sectional analysis of a patient cohort
<sup>c</sup> Lung symptoms selection: no = unselected; required = all RA patients were required to have lung symptoms or suspected ILD; excluded = RA patients with lung symptoms were excluded from recruitment; preRA ex = patients with ILD diagnosed before the RA onset were excluded.
<sup>d</sup> Classification criteria: Multi = multifactorial; CT = chest CT-based; CT/Multi = C-C studies in which the RA-ILD patients were classified with multifactorial criteria and the RA-control with chest CT-based criteria.
<sup>e</sup> Age is the mean current age if available. Otherwise, the median current age is used. Smokers include “ever smokers” and “smokers” in the reports.
<sup>f</sup> Inconsistent information in different parts of the original report. When the difference was large, the data was considered as missing.

Only 13 of the 31 studies reported a significant association of ACPA positivity with RA-ILD. The lack of association could be attributed to insufficient statistical power in some instances but not in others as shown by the very heterogeneous OR, ranging from 0.25 to 42.78. These data confirmed the need for meta-analysis. The total number of patients was sufficient for a sound meta-analysis: 15566 RA patients, divided into 2137 with ILD (8 to 278 per study) and 13429 without ILD (29 to 3491 per study) with few studies missing some ACPA positivity information (six studies missed the ACPA information in 26 to 1486 patients) (Table 1).

### Study design and other characteristics of the data sets

The 31 studies showed very variable designs and attributes that we summarized in a tree focused on the attributes directly related to ILD (Fig. 2). Some aspects were incompletely described requiring the definition of standards (Suppl Methods). The main classes were cohort (CHRT) and case-control (C-C) studies.

**Figure 2:**
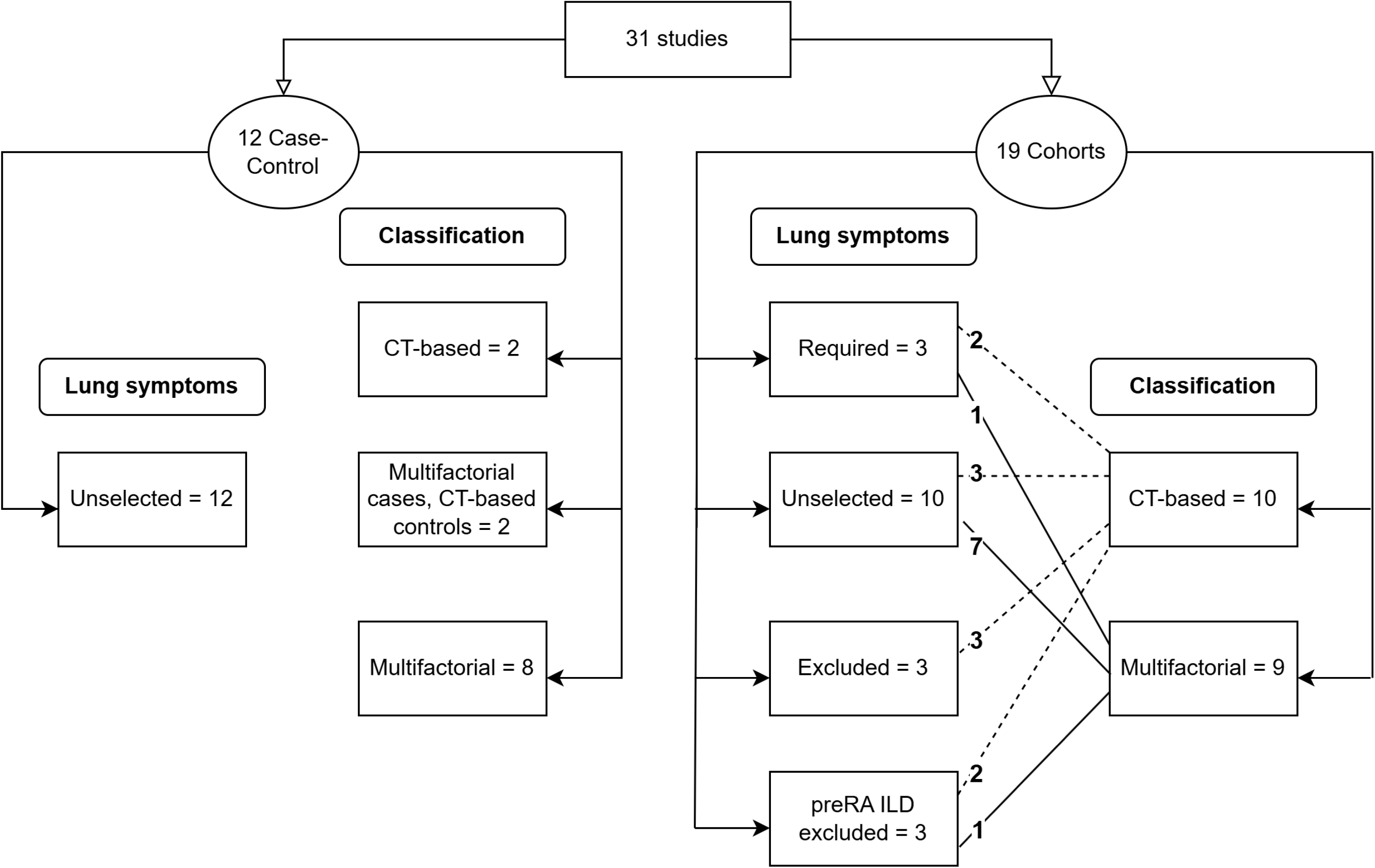
Tree dividing the studies into groups according to the main design characteristics. They were: cohort or case-control; inclusion criteria based on lung symptoms; and patient classification methods.

The 19 CHRT studies were subdivided according to two other characteristics. The first was the presence or absence of an inclusion criterion based on lung symptoms; a criterion used for all patients irrespective of the RA-ILD/RA-control groups. This feature divided CHRT studies into four classes: three studies excluding RA patients with lung symptoms or signs; three studies including only patients with lung symptoms or signs; a third set of three studies excluding patients in whom ILD was already known before RA onset; and the remaining 10 studies without any filter based on lung symptoms (Fig. 2). The second subdivision of CHRT studies was made according to the procedure for patient classification into RA-ILD and RA-control groups. Ten CHRT studies employed chest CT (either standard CT or HRCT); the other nine CHRT studies used multifactorial classification (defined in Suppl. Methods). The two forms of dividing the CHRT studies were correlated: 7/9 studies with any lung inclusion criteria relied on chest CT for classification, whereas only 3/10 studies without a lung symptoms filter did.

None of the 12 C-C studies employed lung symptoms criteria for inclusion of all patients (Fig. 2). The C-C studies differed from CHRT studies also in the use of different classification methods for cases and controls leading to three classes: two studies using CT classification; two studies using CT classification for controls and multifactorial criteria for cases; and the remaining eight studies using multifactorial classification for cases and controls (Fig. 2).

The variability in study designs was compounded by other study characteristics, like recruitment restricted to hospitalized inpatients, participants in tocilizumab randomized clinical trials, or women (Suppl. Table 1). In addition, the patient-level features, such as age, gender, smoking habit, and RF positivity, were also highly variable (Table 1). For example, ever smokers ranged from 8.9 to 79 % or RF positivity from 48 to 98 %. Other variables we extracted from the studies were missing or inconsistently reported in multiple studies, making a meaningful analysis impossible. They included recruitment period, ethnicity, RA duration, age at RA diagnosis, age at ILD diagnosis, ACPA assay, and RA disease activity.

### Baseline heterogeneity, outlier detection, baseline effect, and publication year explorations

Our initial meta-analysis was done to establish the baseline heterogeneity without any moderator. The results showed a significant summary association of ACPA with RA-ILD (OR = 2.32, 95% CI = 1.7-3.1) with a highly significant heterogeneity (p_Q_ = 5.7 x 10^-7^) that was largely attributable to between-study heterogeneity (I^2^ = 62 %). These results were expected given the previously published meta-analyses [12-16]. However, we were surprised to find a study with a disproportionate influence in the summary effect size and heterogeneity (Fig. 3A). The *Kelly, 2014* study alone accounted for an extraordinary fraction (58.1 %) of the between-study heterogeneity and biased all the fitted values on the meta-analysis (Fig. 3B). No other study approached the impact of this one. Also, it showed a large departure from the generality of the studies in our next analysis, a baseline plot (Fig. 3C).

**Figure 3:**
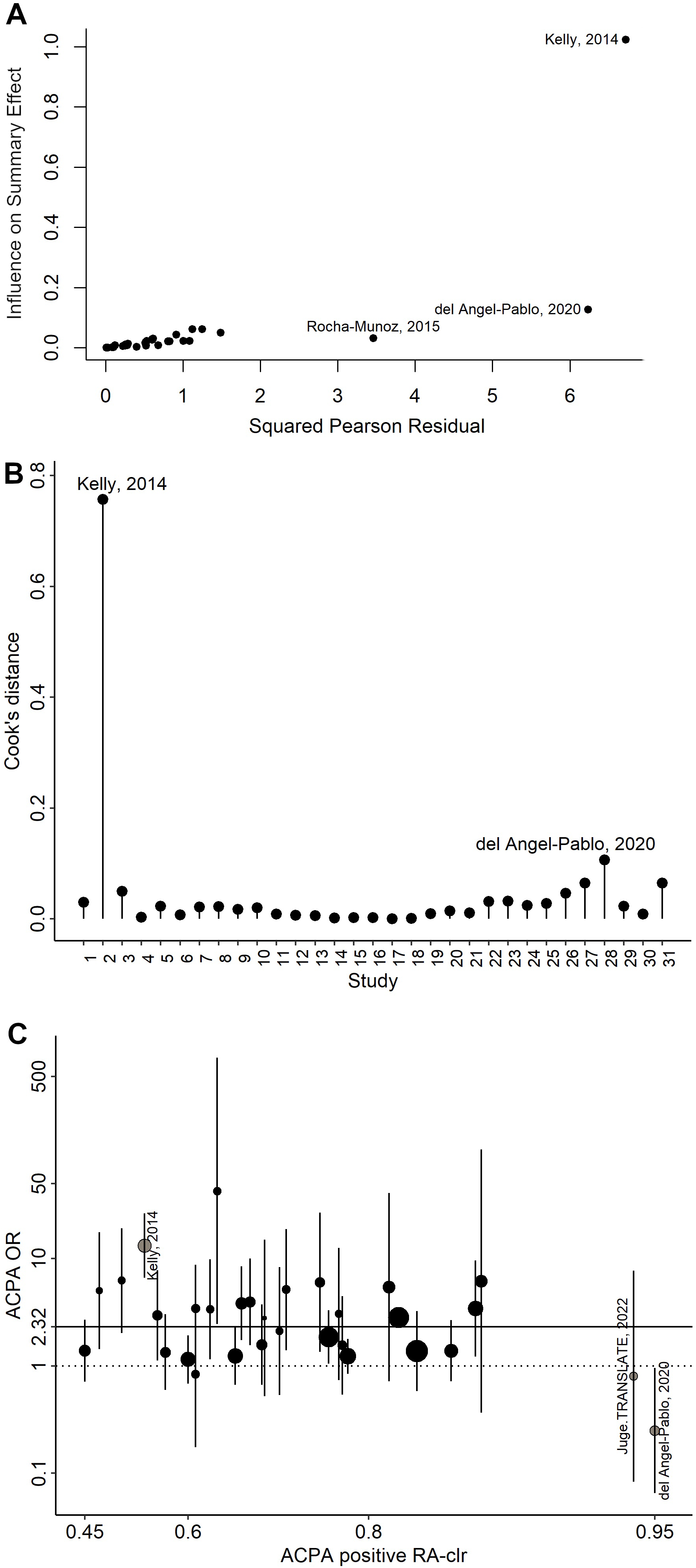
Assessment of the meta-analysis validity. A) Baujal plot showing the impact each study has in the meta-analysis summary effect size (y-axis) and heterogeneity (x-axis). B) Plot of the Cook distances corresponding to each study. This influence measure indicates each study’s impact on all the fitted values in the model. C) Baseline plot showing the distribution of the effect sizes (OR and their CI, y-axis) along the range of ACPA-positive RA-control percentages (x-axis) for each study. The horizontal continuous line represents the summary effect size of the 31 studies (OR = 2.32), whereas the dotted line corresponds to no association of ACPA with RA-ILD (OR = 1). The x-axis is on the natural logarithm scale, the y-axis is the logit scale, and the diameter of the OR circle is proportional to the square root of the number of subjects.

We did the baseline plot to explore if ACPA baseline frequency could contribute to the heterogeneity of the ACPA association with RA-ILD. This type of influence seemed possible due to the wide range of ACPA frequencies in the RA-control group (46.7 to 95.2%). However, the baseline plot showed no OR change in function of the ACPA frequency (Fig. 3C).

As mentioned in the Introduction, we detected a decrease in the summary effect size with the time of publication of previous meta-analyses. Therefore, we conducted a meta-regression analysis taking the time since publication as the moderator (Table 2). The results showed an almost significant (p = 0.061) decrease. This trend required the *Kelly, 2014* study as demonstrated by the nonsignificant meta-regression (p = 0.26) once this study was excluded (Table 2). Therefore, the decrease in the summary effect size of the ACPA association with RA-ILD reflects the published studies but is attributable to the influence of an outlier.

**Table 2.** Initial meta-analysis: outlier impact, time since publication, small study bias, and focus on ACPA.

| Analysis | Studies | Estimate | Estimate *p*-value | *p<sub>Q</sub>*<sup>a</sup> | *I*<sup>2</sup> (95% CI)<sup>a</sup> | $\tau^2$ (95% CI)<sup>a</sup> | % acc.<sup>a,b</sup> |
| --- | --- | --- | --- | --- | --- | --- | --- |
| **Meta-analysis** |  | **OR (95% CI)** |  |  |  |  |  |
|  | All = 31 | 2.32 (1.72, 3.11) | 2.5 x10<sup>-6</sup> | 5.7 x10<sup>-7</sup> | 62 (41, 83) | 0.33 (0.13, 0.94) | ref. |
|  | 30<sup>c</sup> | 2.04 (1.58, 2.64) | 3.39 x10<sup>-6</sup> | 0.004 | 41 (14, 79) | 0.14 (0.03, 0.75) | 58.1 |
| **Meta-regression<sup>d</sup>** |  | **$\beta$ (95% CI)** |  |  |  |  |  |
| Time since publication | All = 31 | 0.11 (-0.005, 0.23) | 0.061 | 3.8 x10<sup>-6</sup> | 59 (35, 80) | 0.28 (0.11, 0.81) | 13.5 |
|  | 30<sup>c</sup> | 0.06 (-0.05, 0.17) | 0.26 | 0.004 | 41 (14, 77) | 0.14 (0.03, 0.70) | 0.0 |
| Study size | 30<sup>c</sup> | 51.6 (4.6, 98.7) | 0.033 | 0.41 | 19 (0, 78) | 0.05 (0, 0.70) | 66.4 |
| ACPA focus | 10 vs. 20<sup>c</sup> | -0.04 (-0.58, 0.50) | 0.88 | 0.003 | 39 (16, 80) | 0.13 (0.04, 0.81) | 1.7 |

<sup>a</sup> Measures of heterogeneity: $p_Q$ = p-value of the overall heterogeneity in meta-analysis or p-value of the residual heterogeneity in meta-regression; $I^2$ = fraction of the heterogeneity attributable to between-study heterogeneity; $\tau^2$ = estimate of the between-study heterogeneity; % acc. = % of the between-study heterogeneity accounted for by the moderator.
<sup>b</sup> The reference is the more general model in each nested series of models.
<sup>c</sup> 30 studies: *Kelly, 2014* was excluded from these analyses. All subsequent analyses and tables excluded this study.
<sup>d</sup> Each meta-regression included a moderator: Time since publication, the squared root of the number of months from publication to bibliographic search; Study size, the inverse of the number of patients with ACPA information; and ACPA focus, the presence (n = 10) or absence (n = 20) of a high focus on ACPA.

The above results motivated a revision of the characteristics of the *Kelly, 2014* study [24]. The study reports the recruitment of RA-ILD patients from multiple centers across the United Kingdom diagnosed over 25 years *versus* RA-controls recruited from a single center at the time of the analysis. In addition, 26.1 % of the RA-ILD patients missed ACPA information but none of the controls. Considering its significant influence and differentiated recruitment of cases and controls, we decided to exclude the *Kelly, 2014* study from subsequent analyses (Table 2). This measure resulted in a reduced association of ACPA with RA-ILD (OR = 2.04, 95% CI = 1.6-2.6) and a much lower between-study heterogeneity (p_Q_ = 0.004).

### Detection of small study bias in the remaining 30 studies

Another exploration step found significant evidence of small study bias in the funnel plot (Sup. Fig. 1, Egger’s test p = 0.004 and Rank test p = 0.038). The stronger association between ACPA and RA-ILD in small studies than in large studies was confirmed by meta-regression with the study size as moderator (Table 2). This finding suggests publication bias but could have other causes. To assess the likelihood of publication bias, we examined if the bias correlated with the importance attributed to ACPA in each study. This analysis divided the studies into two classes: 10 studies with a focus on ACPA because they mentioned ACPA in the article title, the ACPA assay was done specifically for the study, or the ACPA analysis was a stated study goal; and the other 20 studies without a focus on ACPA. The meta-regression analysis did not show evidence of a stronger association of ACPA with RA-ILD in the studies with a focus than in those without it (p = 0.88, Table 2), questioning that the small study bias could be attributed to publication bias.

### The patient classification method as a significant moderator

Next, we explored the influence of the study design (Table 3). The major division was between CHRT and C-C studies (Fig. 2). However, this design factor did not determine significant differences in the ACPA association with RA-ILD (p = 0.83, Table 3). We continued exploring strata within the CHRT studies starting with the four classes based on the use of lung symptoms for inclusion (Fig. 2): symptoms required for inclusion, symptomatic patients excluded, patients with ILD preceding RA excluded, or lung symptoms not considered. The meta-regression showed no significant impact of this characteristic (Table 3). The result was consistently nonsignificant in a multinominal analysis of the four classes and the three separate analyses of studies with each lung inclusion criteria compared to studies without this type of inclusion criteria (Table 3). Another design factor within the CHRT studies was the patient classification method (Fig. 2): chest CT or multifactorial criteria. The meta-regression showed a significantly stronger association of ACPA with RA-ILD in the CT group than in the multifactorial criteria studies (p_βclas._ = 0.0048, Table 3). This significant moderator explained a large fraction of the between-study heterogeneity (98.7%) leading to the nonsignificance of the heterogeneity test (p_Q_ = 0.25). The subgroup meta-analysis produced estimates of the association effect sizes in each study class: OR = 2.80 in the CT *vs.* OR = 1.47 in the multifactorial criteria studies.

**Table 3.**
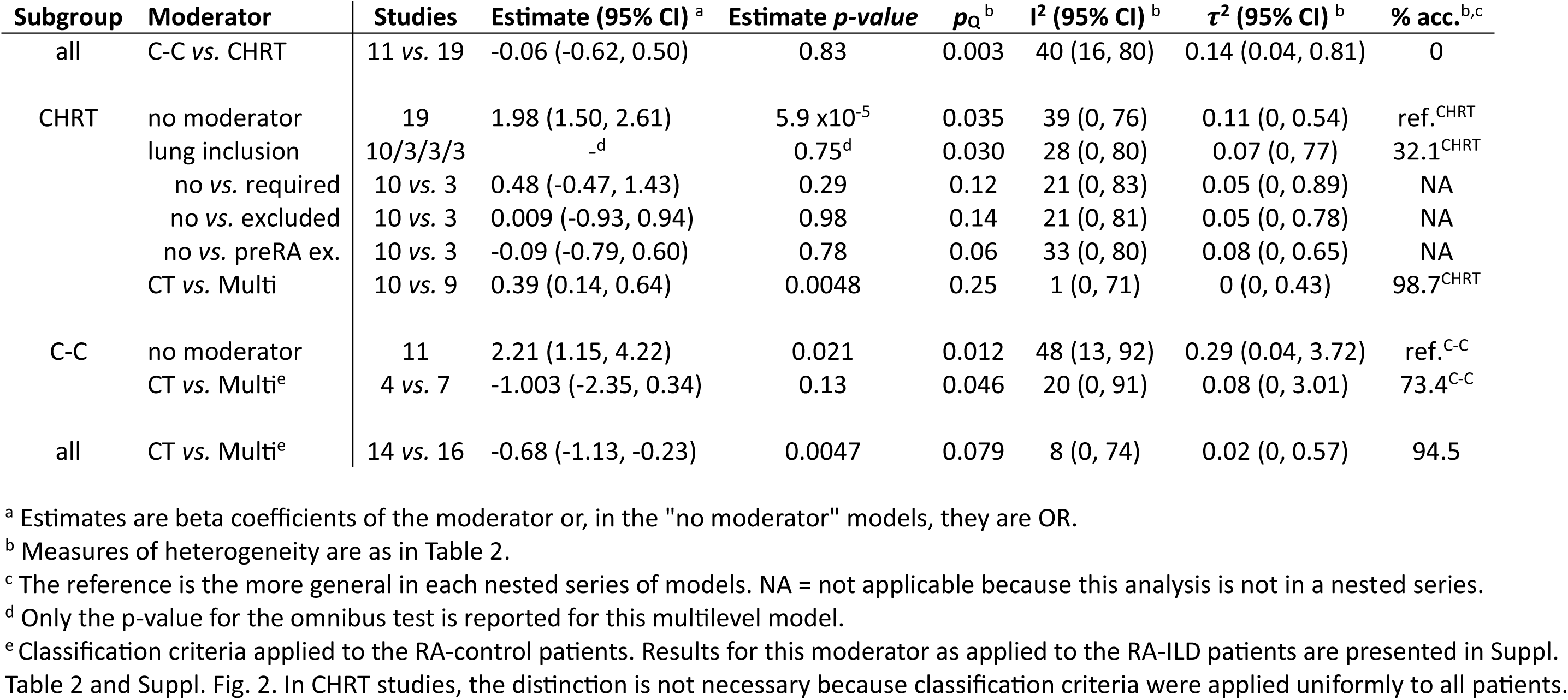
Meta-regression analyses including study design characteristics: CHRT/C-C; lung selection criteria; and CT-based/Multifactorial classification criteria.

| Subgroup | Moderator | Studies | Estimate (95% CI)<sup>a</sup> | Estimate *p*-value | *p*<sub>Q</sub><sup>b</sup> | I<sup>2</sup> (95% CI)<sup>b</sup> | τ<sup>2</sup> (95% CI)<sup>b</sup> | % acc.<sup>b,c</sup> |
| --- | --- | --- | --- | --- | --- | --- | --- | --- |
| all | C-C vs. CHRT | 11 vs. 19 | -0.06 (-0.62, 0.50) | 0.83 | 0.003 | 40 (16, 80) | 0.14 (0.04, 0.81) | 0 |
| CHRT | no moderator | 19 | 1.98 (1.50, 2.61) | 5.9 x10<sup>-5</sup> | 0.035 | 39 (0, 76) | 0.11 (0, 0.54) | ref.<sup>CHRT</sup> |
|  | lung inclusion | 10/3/3/3 | -<sup>d</sup> | 0.75<sup>d</sup> | 0.030 | 28 (0, 80) | 0.07 (0, 77) | 32.1<sup>CHRT</sup> |
|  | no vs. required | 10 vs. 3 | 0.48 (-0.47, 1.43) | 0.29 | 0.12 | 21 (0, 83) | 0.05 (0, 0.89) | NA |
|  | no vs. excluded | 10 vs. 3 | 0.009 (-0.93, 0.94) | 0.98 | 0.14 | 21 (0, 81) | 0.05 (0, 0.78) | NA |
|  | no vs. preRA ex. | 10 vs. 3 | -0.09 (-0.79, 0.60) | 0.78 | 0.06 | 33 (0, 80) | 0.08 (0, 0.65) | NA |
|  | CT vs. Multi | 10 vs. 9 | 0.39 (0.14, 0.64) | 0.0048 | 0.25 | 1 (0, 71) | 0 (0, 0.43) | 98.7<sup>CHRT</sup> |
| C-C | no moderator | 11 | 2.21 (1.15, 4.22) | 0.021 | 0.012 | 48 (13, 92) | 0.29 (0.04, 3.72) | ref.<sup>C-C</sup> |
|  | CT vs. Multi<sup>e</sup> | 4 vs. 7 | -1.003 (-2.35, 0.34) | 0.13 | 0.046 | 20 (0, 91) | 0.08 (0, 3.01) | 73.4<sup>C-C</sup> |
| all | CT vs. Multi<sup>e</sup> | 14 vs. 16 | -0.68 (-1.13, -0.23) | 0.0047 | 0.079 | 8 (0, 74) | 0.02 (0, 0.57) | 94.5 |

<sup>a</sup> Estimates are beta coefficients of the moderator or, in the "no moderator" models, they are OR.
<sup>b</sup> Measures of heterogeneity are as in Table 2.
<sup>c</sup> The reference is the more general in each nested series of models. NA = not applicable because this analysis is not in a nested series.
<sup>d</sup> Only the *p*-value for the omnibus test is reported for this multilevel model.
<sup>e</sup> Classification criteria applied to the RA-control patients. Results for this moderator as applied to the RA-ILD patients are presented in Suppl. Table 2 and Suppl. Fig. 2. In CHRT studies, the distinction is not necessary because classification criteria were applied uniformly to all patients.

The C-C studies were uniform in not using lung symptoms for inclusion. However, they were more varied than CHRT studies in the patient classification procedures including a class with different methods in RA-ILD and RA-controls (Fig. 2). Therefore, we performed two lines of analysis, one according to the classification method used for the RA-ILD group and the other according to the one used for RA-controls. The RA-control analysis is presented in detail, whereas the RA-ILD analysis is shown only as part of the sensitivity analysis because the two produced similar results. In the C-C studies, the classification procedure did not significantly modify the ACPA association with RA-ILD (Table 3). However, this result can be attributed to the small number of studies because, like in the CHRT studies, the OR was nominally larger in the CT-based studies (OR = 4.58) than in the multifactorial criteria studies (OR = 1.68). The combined analysis of CHRT and C-C studies (Fig. 4 and Suppl. Fig. 2) ratified the stronger ACPA association with RA-ILD in the CT than in the multifactorial criteria group (OR = 3.05 *vs.* OR = 1.55, p_βclas._ = 0.0047). Notably, the classification method explained a large fraction of the between-study heterogeneity (94.5 %) resulting in nonsignificant residual heterogeneity (p_Q_ = 0.079) also in the analysis of the 30 studies (Table 3).

**Figure 4:**
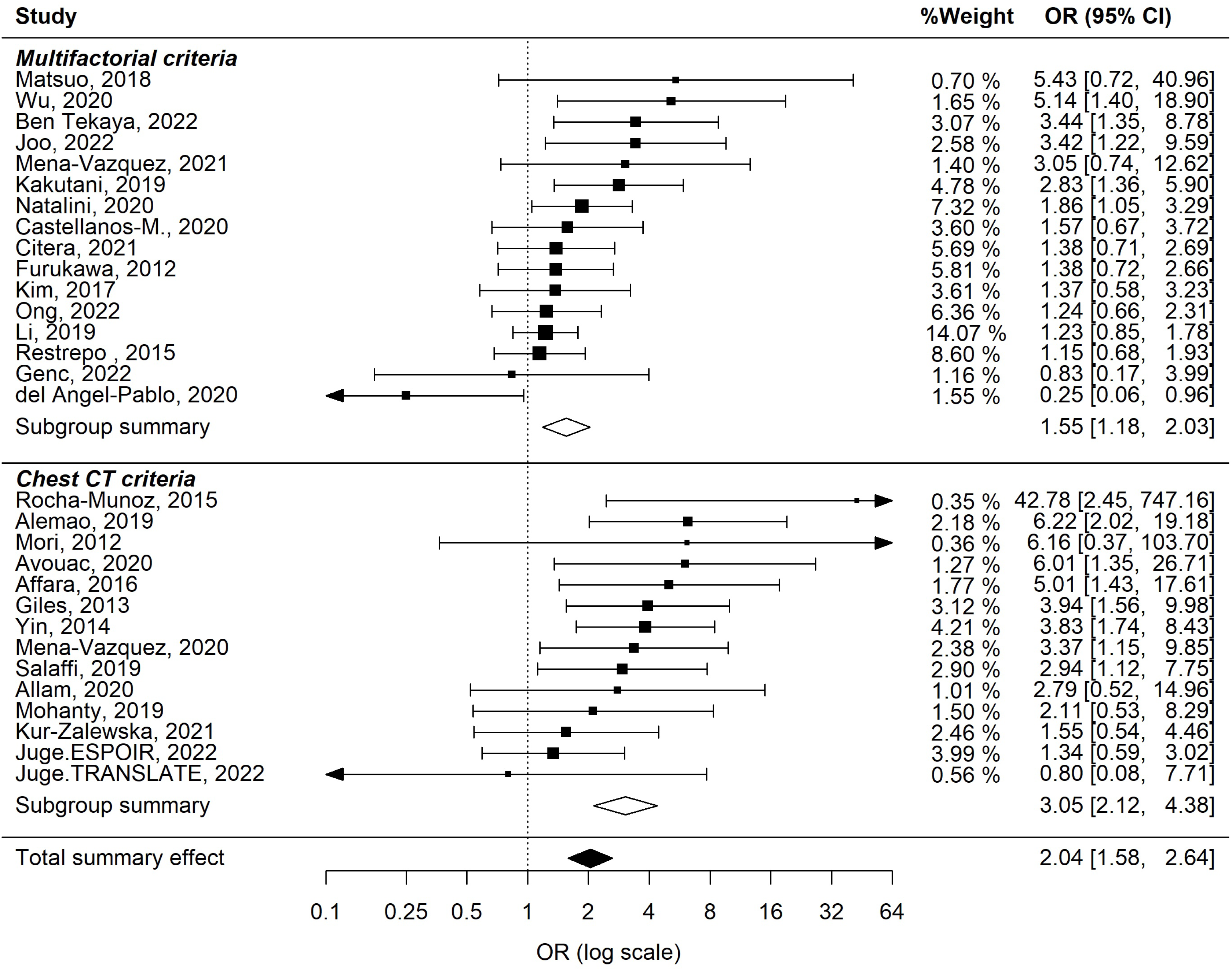
Subgroup meta-analysis of ACPA association with RA-ILD according to the patient classification method. This plot shows the classification method for the RA-control group. A similar plot for the RA-ILD subgroup is shown in Suppl. Fig. 2. The squares and bars represent OR and 95 % CI, respectively, which are reported in the rightmost column. The size of the squares is proportional to the weight of each study, which is also reported in the first right column. Subgroup and overall summary effects sizes are represented by diamonds whose longest axis is the 95% CI. Heterogeneity measures are reported in Table 3.

The sensitivity analyses showed the patient classification method was a robust moderator of the ACPA association (Suppl. Table 2). These meta-regression analyses included modifications to technical aspects of the meta-regression: changing Knapp and Hartung test adjustment by the Z distribution (p_βclas._ = 0.00045); replacing Maximum Likelihood by Restricted Maximum Likelihood (p_βclas._ = 0.0011); the full range of assumptions behind the analysis from fixed-effect (p_βclas._ = 0.0002) to equal-weight meta-regression (p_βclas._ = 0.021); the exclusion of studies with > 5% missing ACPA information (p_βclas._ = 0.010); and the exclusion of one of each pair of articles with potentially overlapping data (p_βclas._ = 0.0091 or 0.019, depending on the excluded report). Other sensitivity analyses showed the classification method was also robust to modifications in important study-level features (Suppl. Table 2): analysis restricted to the 26 studies with an unambiguous distinction between CT and multifactorial criteria (p_βclas._ = 0.017), to the studies that specifically used HRCT for CT classification (p_βclas._ = 0.022); and when the distinction between CT and multifactorial criteria was based in the RA-ILD group (p_βclas._ = 0.013).

### No other moderators except the effect of age and RF conditional on the classification method

None of the other study characteristics was a moderator of the ACPA association with RA-ILD (Suppl. Table 3). The analyzed characteristics included a variety of features related to the studies and others at the patient level. Regarding the study-level characteristics, we analyzed: location, monocentric or multicentric nature, allowance for concomitant systemic autoimmune diseases, restriction to hospitalized inpatients or patients treated with specific drugs, a high bias for one sex or young patients, and the use of CT or HRCT in patient assessment (Suppl. Table 3). In addition to these features, we analyzed four patient-level characteristics: current age, sex, smoking habit, positivity for rheumatoid factor, and the fraction of patients with RA-ILD. The first three were considered for all patients, the RA-ILD, the RA-controls, and the difference between RA-ILD and RA-controls (Suppl. Table 3).

The same study characteristics were assessed for their potential effect conditional on the patient classification using two-factor meta-regression analyses (Suppl. Table 4). These analyses were nonsignificant except for two patient-level characteristics: current age and the difference in RF frequency between RA-ILD and RA-controls (in other words, the RF association with RA-ILD). The first conditional effect was observed with the age of the RA-controls (p = 0.047) or the whole study (p = 0.032), but not with the age of the RA-ILD patients or the difference in age between RA-ILD and RA-controls. The conditional effect consisted in the weaker association of ACPA with RA-ILD in the studies with younger RA-controls (or RA patients overall) only in the studies relying on multifactorial criteria (Fig. 5A). In contrast, the ACPA association was unmodified by age in the studies employing CT classification. The inclusion of age in the meta-regression showed a significantly better fit to the data than without age, as shown with the AIC, BIC, and LRT analyses (Suppl. Table 4). Regarding the RF, the two-factor meta-regression revealed a positive correlation between the RF and the ACPA associations with RA-ILD in each of the two types of study (p = 0.0090; Fig. 5B). This effect was nonsignificant without the separation into CT and multifactorial studies (p = 0.091; Suppl. Table 3). The two-factor meta-regression fitted the data significantly better than without the RF association factor (Suppl. Table 4).

**Figure 5:**
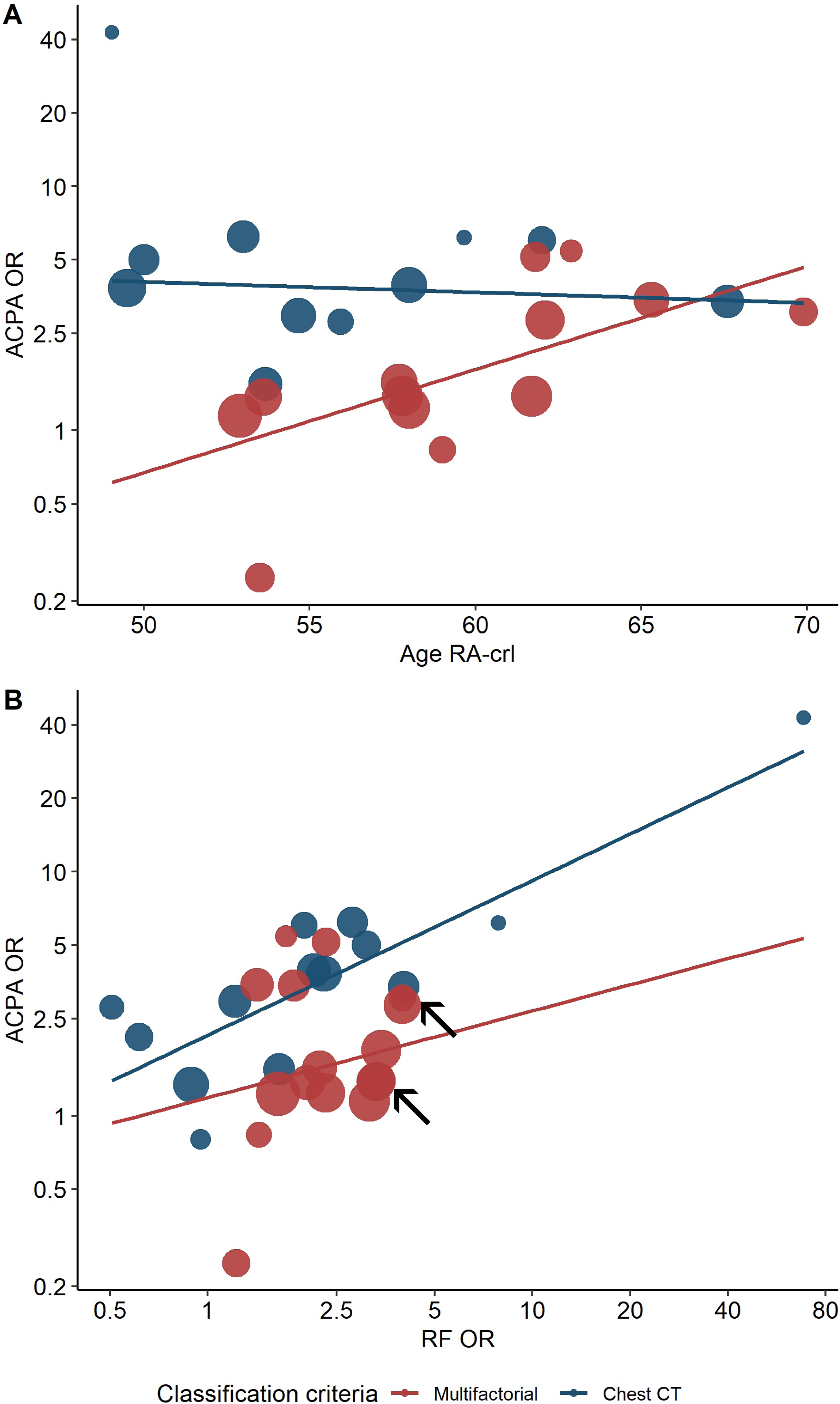
Two patient-level features affect the ACPA association with RA-ILD conditional on the classification method. Bubble plots showing the conditional effects of A) the current age of the RA-control group; and B) the RF association with RA-ILD. The color of the bubbles identifies the studies according to the chest CT (blue)/Multifactorial (red) classification method. The size of the bubbles is proportional to the weight of the study in the random effect meta-regression. The two arrows signal bubbles where two multifactorial classified studies overlap almost completely. The y-axis shows the studies OR (in the logarithm scale) and the x-axis represents in A) the current age of RA-control (in years); in B) the OR of the association of RF positivity with RA-ILD (in the logarithm scale).

Finally, we observed that CT classification was more commonly used in small studies than in large ones. Therefore, we wondered if the classification procedure could account for the small sample bias we had detected in the initial analyses. The two-factor meta-regression (study size and classification method) was consistent with this hypothesis because the study size lost its significant effect on the ACPA association (p = 0.38, Suppl. Table 5) but inconclusive because the classification procedure only retained a non-significant trend (p = 0.058).

## Discussion

Our exploration has uncovered that an outlier with differentiated case-control recruitments and the patient classification method account for the heterogeneity of ACPA association with RA-ILD in the bibliography. These results highlight the importance of study design and the dependence of strong ACPA association on the patient classification using chest CT. The remaining heterogeneity was nonsignificant, meaning that is undistinguishable from random variation. Additionally, our results validate the ACPA association with RA-ILD, providing arguments in favor of the assessment of ACPA in the screening strategies for RA-ILD.

The discovery of the disproportionate influence of *Kelly, 2014* has been key to obtaining results that are not unduly biased. This study pushed the heterogeneity of the meta-analysis to a very significant level (from p_Q_ = 0.004 to 5.7 x10^-7^), an extraordinary increase for a single study. Our review found notable differences in the time and place of recruitment between cases and controls in that study, which could explain its outlier behavior [24]. The two elements justified our decision to exclude *Kelly, 2014* from the analysis.

The moderator effect of the patient classification method was observed considering the two groups: RA-ILD and RA-control. The analyses considering each group overlapped extensively because only two studies employed a different classification method for each group. Therefore, it was impossible to distinguish between them. This circumstance complicates the interpretation of the findings because we are forced to consider the two patient groups. The two differ in many aspects including the unequal assessment in the studies using multifactorial criteria. In the RA-control group, the multifactorial procedure could be as undiscriminating as the absence of a diagnosis of ILD. In contrast, the multifactorial criteria in the RA-ILD group commonly included chest CT in addition to the presence of symptoms and other explorations. Therefore, we are inclined to attribute the moderator effect to the improved classification of the RA-control group obtained with systematic CT exams. This method uncovers unsuspected RA-ILD patients. These patients would be false RA-controls in the absence of chest CT and decrease the ACPA association with RA-ILD. However, we do not need to distinguish between the impact of classification methods in the RA-ILD and RA-control groups for a more general interpretation of the findings: the ACPA association with RA-ILD would be more directly related to the presence or absence of ILD lesions in the chest CT than to ILD clinical manifestations.

The sensitivity analyses validated the significant moderator effect of the patient classification methods. These analyses included multiple aspects of potential influence. Two deserve commentary because they affect the classification methods themselves. One is the ambiguity about the method used in some articles. We verified that the exclusion of the ambiguous studies did not cancel the moderator effect. However, it will be better to know this important information for all studies without doubts, stressing the need for reporting sufficient information on the design and methods in each publication. Another aspect concerns our decision to consider chest CT as encompassing standard CT and HRCT. We addressed this aspect in three analyses, a sensitivity analysis, a potential moderator analysis, and an analysis for conditional factors, but none of these analyses suggested the need to separate the four standard CT studies from the HRCT ones.

Our analyses also uncovered revealing effects of two patient features conditional on the classification method. The age conditional effect revealed a larger impact of the classification method in studies with younger patients: it was in these studies where the difference in the ACPA association with RA-ILD between the CT and multifactorial criteria was largest. This result can be interpreted as reflecting an increased frequency of unsuspected ILD in the younger RA-control group, which is likely because older age is associated with ILD [1, 2, 10]. Therefore, it is possible to hypothesize that older groups of RA-controls would contain fewer false RA-control patients despite the absence of chest CT. In turn, the conditional effect of the RF association revealed the correlation between the ACPA and RF associations with RA-ILD in each of the subgroups defined by the patient classification method. This result is consistent with the well-known positive correlation between the two RA autoantibodies, as they are commonly found to be present or absent together [25-27]. In addition, multiple pieces of evidence indicate that the concomitant presence of the two antibodies potentiates their contribution to RA pathogenesis, including RA risk, inflammation, and bone erosions [27-31].

Our results clarify the inconsistent ACPA association with RA-ILD in the analyzed studies. It was due to deficiencies in the design of the studies and limitations of the patient classification, not to lack of association. In addition, the association was of a sufficient magnitude in the studies using chest CT (OR = 3.05) to be of potential utility. Whether it is useful as part of a screening score cannot be answered with the studies reviewed here; this question requires studies specifically designed to define screening instruments [3, 32]. However, the wide availability of ACPA and the preferential association when the RA-ILD status is defined with chest CT go in the right direction because the screening aim is to identify patients requiring chest CT and intensive follow-up [2-5].

Additionally, our findings strengthen the ACPA involvement in RA-ILD pathogenesis [1, 33]. This involvement is suggested by several pieces of evidence, like the presence of B cell follicles containing germinal centers and positive for ACPA staining in the lung [34, 35], or the enrichment of ACPA in the bronchoalveolar lavage and the induced sputum from RA patients [36, 37]. These findings likely reflect the local ACPA production in the lungs of patients with RA, but none of them established the link with ILD as they were obtained in patients with RA, without being selected for RA-ILD. Only the epidemiological demonstration of increased RA-ILD risk in ACPA-positive patients bridges this gap in the pathogenic hypothesis [1, 33].

None of the other study-level or patient-level characteristics was a significant moderator of the ACPA association with RA-ILD. This negative result does not exclude the influence of characteristics that were present in a few studies. They could have escaped detection for lack of statistical power. We think this limitation could apply for example to the inclusion criteria based on lung symptoms because each class of criteria was present only in three studies. Additionally, some characteristics of interest were not analyzed due to the lack of appropriate data. One was ethnicity, which was not reported or lacked the ACPA frequencies for each ethnic group when more than one was studied. We replaced ethnicity with the study location, which was not a significant moderator. Other moderators we could not analyze are age at RA or ILD diagnosis and RA duration, which were not reported or inconsistently: mean, median, or maximum duration.

## Conclusions

In conclusion, our exploration has been successful in identifying two sources of heterogeneity in the reports of ACPA association with RA-ILD: an outlier study and the patient classification method. The two highlight the importance of the study design. Once the two sources were accounted for, the heterogeneity was not significantly different from random variation. These findings support the consideration of ACPA as a faithful RA-ILD biomarker. However, its value for the screening of RA-ILD requires another type of study. Additionally, the discovery of a stronger association in the studies applying chest CT for classification suggests the possibility it could be an efficient approach in the search for other RA-ILD biomarkers.

## Data Availability

All data produced in the present work are contained in the manuscript and supplementary material

## List of abbreviations

ACPA: anti-citrullinated protein antibodies
ILD: interstitial lung disease
MUC5B: mucin 5B gene
OR: odds ratio
RA: rheumatoid arthritis
RA-ILD: rheumatoid arthritis-associated interstitial lung disease
AIC: Akaike information criterion
BIC: Bayesian information criterion
C-C: case-control
CHRT: cohort
CI: confidence interval
CT: computed tomography
HRCT: high-resolution computed tomography
LRT: likelihood ratio test
RF: rheumatoid factor

## Declarations

### Ethics approval and consent to participate

Not applicable

### Consent for publication

Not applicable

### Availability of data and materials

All data analyzed during this study are included in this published article and its supplementary information files

### Authors’ contributions

BK contributed to study conceptualization and design; data extraction and curation; formal analysis, interpretation of the results, figure design, writing the original draft, revision, edition, and final manuscript approval. CC participated in study design; data extraction and curation; interpretation of the results, revision, edition, and approval of the final manuscript. AG contributed to study conceptualization and design; supervision, funding, and project management; formal analysis, interpretation of the results, writing the original draft, revision, edition, and final manuscript approval.

## Acknowledgments

The MARILD network members are acknowledged for their encouragement and support towards a better understanding of RA-ILD.

