## Supplementary text, figures and tables for "Meta-analysis identifies the patient classification major impact on the ACPA association with rheumatoid arthritis-associated interstitial lung disease (RA-ILD)"

### Supplementary Methods

#### Article selection

Article selection, classification, and data extraction were performed independently by two researchers (BK and CC) following the PRISMA 2020 recommendations (1). All the discrepancies between the reviewers were shared and resolved by consensus. The study was not preregistered. Only original research publications written in English that included the frequencies of ACPA-positive RA-ILD and RA-control patients were selected. Analysis of the frequency of ACPA did not have to be the main study objective. Publications with duplicated information were excluded, but those with a suspected overlap of patients were retained and subjected to posterior assessment by alternatively excluding one of the potentially overlapping studies, either the most recent or the oldest. All study designs were accepted except for the longitudinal studies reporting only ACPA association with RA-ILD as a function of time (hazard risks). No quality criteria were used for the selection or weighting of the studies.

#### Data extraction

All aspects of the study design were retrieved and tabulated. They included publication date, period of patient recruitment, location, pairing of RA-ILD and RA-control populations, ethnicity, type of study, inclusion and exclusion criteria, RA-ILD and RA-control classification method, fraction of RA-ILD patients, RA duration, ACPA assay, ACPA association, and place of ACPA on the study. Also, we noted every peculiarity of the studies that could be interesting. Examples were studies that included only hospitalized patients, did not exclude patients with overlapping systemic autoimmune disease, or selecting their patients among participants in clinical trials. Other variables previously related to RA-ILD risk, such as age (at RA onset, ILD diagnosis, and current age), sex, smoking status, rheumatoid factor positivity, disease activity and BMI were also extracted if possible. These latter variables were retrieved for the RA-ILD and RA-control groups in addition to the whole patients set. Quantitative variables were extracted as means or, secondarily as medians, when available. Frequencies and Odds Ratios (OR) were extracted from the text or calculated based on the data. When the reported OR was adjusted by other factors, we recalculated the OR from the original data without adjustment.

**Study Design factors and their Definitions**

The variables of study design included design as a case-control or cohort study, the selection of patients according to the presence or absence of lung symptoms, and the reliance on chest CT scans or multifactorial criteria for classification among others. To avoid ambiguity about some aspects of the study design, we defined the classes that could be used. These definitions were modified in function of the information we found in the selected studies. However, they were applied uniformly to all studies with reassessment when necessary.

**Cohort studies (CHRT):** those beginning from a collection of patients with RA and searching for ILD among them, recruiting patients independently of their ILD status, all sharing the same place and time of recruitment, and in which the recruitment did not modify the ratio of patients with/without ILD. Contradiction of any of these points led to the classification of the study as a C-C. Some CHRT studies included longitudinal and cross-sectional data, but only the latter was extracted for meta-analysis.

**Case-control studies (C-C):** those in which the two groups, RA-ILD and RA-control, were already known before the study (even if some patients were reclassified during the assessment for study inclusion), they were not recruited from the same place, time, or with compatible criteria, or the ratio of RA-ILD/RA-control cannot be considered representative of the population. Any of these characteristics implied classifying the study as C-C.

**Lung symptoms or signs for inclusion:** studies that selected all the RA patients, irrespective of the RA-ILD/RA-control status, according to the presence of lung symptoms or signs. This selection led to four types of studies:

- **Required:** The presence of lung symptoms or signs was required for inclusion. These studies could be considered as focusing on patients with a suspicion of ILD who after a complete examination were classified as clinical RA-ILD or RA-controls.

- **Excluded:** The presence of lung symptoms or signs was sufficient to exclude from the study. These studies performed chest HRCT on all patients to classify them either as subclinical RA-ILD or ILD-free RA patients. Two C-C studies (*Rocha-Muñoz, 2015* and *Kelly, 2014*) considered an exclusion criterion for being an RA-control the presence of lung symptoms but not for being an RA-ILD case, therefore they were not considered in this class but as unselected.
- **preRA ILD excluded:** The presence of a diagnosis of ILD before the RA diagnosis was sufficient to exclude from the study. Commonly, these studies aimed to identify risk factors with utility to stratify the RA patients according to their RA-ILD risk.
- **Unselected:** The study did not use lung symptoms or signs as selection criteria for patient inclusion.

**Classification method:** The procedures used to classify the patients into RA-ILD and RA-control groups, which we divided into two groups:

- **chest CT:** These studies used only chest CT to ascribe the patients to the RA-ILD or RA-control groups. The CT exams were often HRCT, but they can be also standard CT (Suppl. Table 1).
- **Multifactorial criteria:** This group of studies includes studies that explicitly stated the multifactorial criteria and others that were less specific. Therefore, this definition is used to encompass multifactorial, clinical and conventional criteria. In addition, the multifactorial criteria in the C-C studies were unequal for the cases and the controls: in the RA-ILD group they commonly involved CT, presence of symptoms, and other explorations; in the RA-control group none of these components was necessary.

Additionally, the classification method for RA-ILD and RA-control groups differed in two C-C studies. In these two studies, the RA-ILD patients were classified using multifactorial criteria and the RA-controls were included after a chest CT. Besides, there were six studies where the distinction between CT and multifactorial classification could not be made confidently (Suppl. Table 1). These studies were excluded in one of the sensitivity analyses.

**Detailed meta-analysis and meta-regression methodology to identify the causes of heterogeneity**

The use of meta-analysis, meta-regression and subgroup meta-analysis for the investigation of heterogeneity between studies is well developed although less well-known than the use of meta-analysis to summarize data (2-7). The summary and meta-regression effects were estimated with random effect meta-analysis employing the inverse variance method for the studies' weights and maximum likelihood for estimating the between-study variance (tau-squared, $\tau 2$). The maximum likelihood estimation is appropriate for meta-analysis focused on between-study heterogeneity because it allows tests for model selection (6). These tests include the Akaike Information Criterion (AIC), Bayesian Information Criterion (BIC), and the likelihood ratio test (LRT). All the model comparisons were made on nested models: the model with more moderators was compared with an identical model except that it contained fewer moderators (6). The Knapp and Hartung test adjustment for the p values was used (3, 6). Conditional analysis was performed with two-factor meta-regression without interaction terms. No higher-order conditional or interaction models were explored to minimize the risk of artifacts caused by ecological bias, the identification of spurious moderators because the factors co-aggregate across studies (3). The significant moderators were subjected to sensitivity analysis across a set of variations in the meta-analysis methodology and other factors of the studies or variable definitions that could cause artifactual results. One of the meta-analysis assumptions tested was the model for the between-studies heterogeneity: changing the random effects model to a fixed effect model at one extreme, and to a model of infinite heterogeneity at the other (8).

Assessment of the meta-analysis included a search for outliers, publication bias, and baseline effects (3, 6, 9). A series of influence measures included in the *metafor* package were used to identify potential outliers (6, 9). Three were given special attention: squared Pearson residual (it quantifies the contribution of the study to the Q statistic, the parameter measuring overall heterogeneity), influence on the fitted value (it measures the study's impact on the summary effect size), and Cook's distance (a distance representing the change in all the fitted values in the model). The two first are used as the x and y axes of the Baujal plot. The three influence measures are obtained by comparing the meta-analysis with all the studies and the recalculated analyses after excluding each study. The publication bias was assessed with the funnel plot and tests for the asymmetry of the distribution of studies' effect sizes relative to their standard errors (6). The presence of a baseline effect on the ACPA association with RA-ILD was explored with the baseline plot as we find it easier to interpret than the L'Abbé plot (3, 10). These plots check if the effect sizes depend on the studies' population positivity rate (the baseline effect). The frequency of ACPA in RA-control patients was used as a surrogate of the population rate.

The effect size of the APCA association with RA-ILD was measured as the natural logarithm of the OR. However, the results are presented after back-transformation to OR for easy interpretation. Potential moderators were the study-level variables described in the previous section and the main manuscript. The patient-level variables were age, and the percentages according to sex, smoking status, and rheumatoid factor positivity. All the qualitative variables were treated as dichotomous. The quantitative variables showing a skewed distribution were transformed: the percentages were logit transformed; time since the publication was the squared root of the number of months; and the size of the study was the inverse of the number of subjects. The localization of the study was treated as two separate factors, longitude and latitude. The coordinates were normalized relative to the center of the distribution. For every moderator, we assessed the regression coefficient, its p-value, and three measures of heterogeneity, Q, $I^2$, and $\tau^2$ (2, 3, 6, 7). Cochrane's Q assesses the overall heterogeneity in the model; the inconsistency index $I^2$ estimates the fraction of heterogeneity attributable to between-study heterogeneity; and $\tau^2$ is the estimated between-study heterogeneity. When qualitative moderators were significant, we also conducted subgroup analysis to obtain the parameters for each stratum. The meta-analysis results are shown as forest plots, whereas the meta-regression results are presented as bubble plots. Meta-analysis and meta-regression were done in JASP (v0.18.3) based on the *metafor* R package (6). All other analyses were done in R (v4.3.2). P-values $< 0.05$ were considered statistically significant.

**Suppl. Fig. 1: Funnel plot of the 30 studies included in the meta-analysis (without *Kelly, 2014*).** It represents the distribution of the studies according to their effect sizes = lnOR (x-axis) and standard errors (y-axis). The colors represent the 90, 95 and 99 % CI. The distribution showed a significant bias.

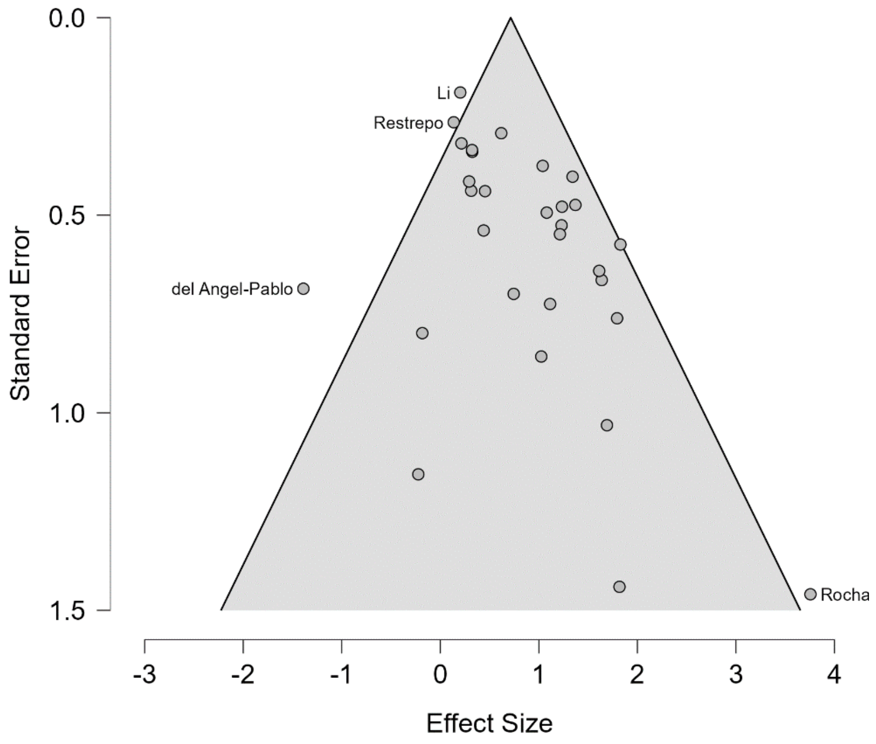

**Suppl. Fig. 2: Meta-analysis of ACPA association with RA-ILD stratified in two subgroups according to the multifactorial or chest CT classification applied to the RA-ILD patients.** It is similar in all respects to Fig. 3 but for the RA-ILD patients (there were two C-C studies with CT-based classification for the RA-control patients but not for the RA-ILD patients).

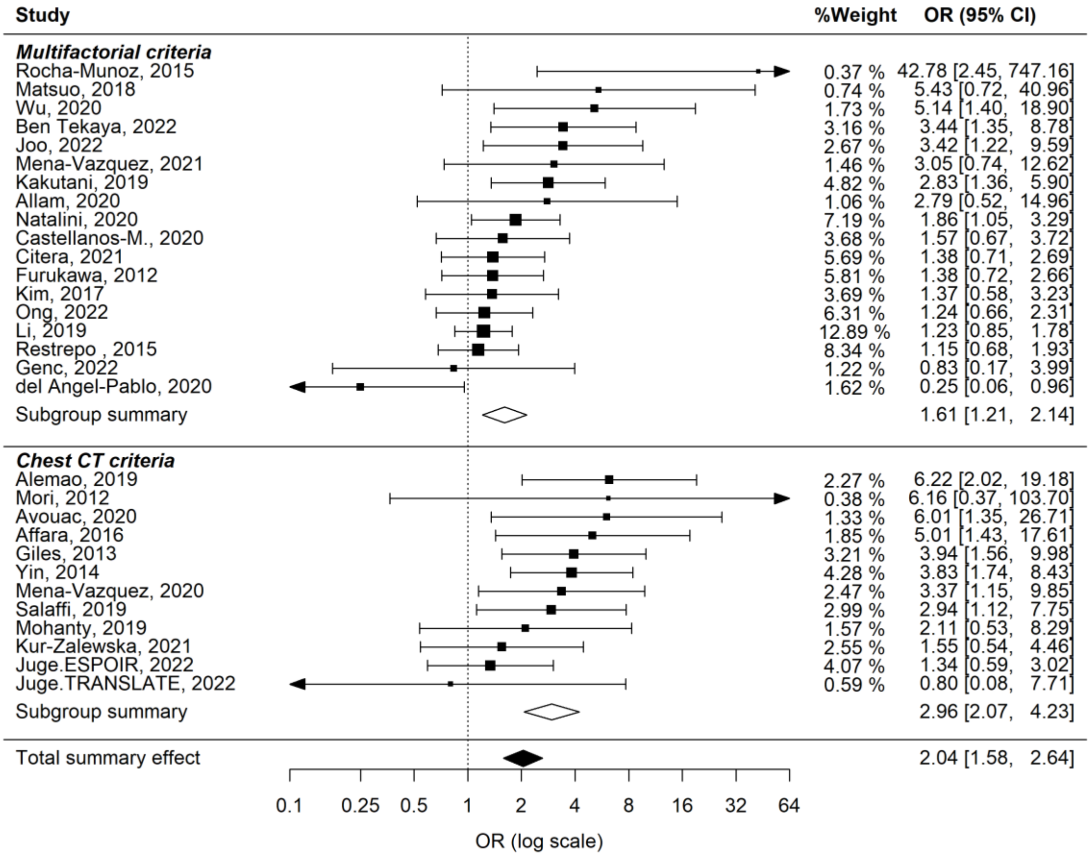

**Supplementary Figure 3: Significant effect of two patient-level features over the ACPA association with RA-ILD conditional on the classification method.** Bubble plots showing the conditional effects of A) the current age of the RA-control group; and B) the RF association with RA-ILD. The color of the bubbles identifies the studies according to the chest CT (blue)/Multifactorial (red) classification method. The size of the bubbles is proportional to the weight of the study in the random effect meta-regression. The two arrows signal bubbles where two multifactorial classified studies overlap almost completely. The y-axis shows the studies OR (in the logarithm scale) and the x-axis represents in A) the current age of RA-control (in years); in B) the OR of the association of RF positivity with RA-ILD (in the logarithm scale).

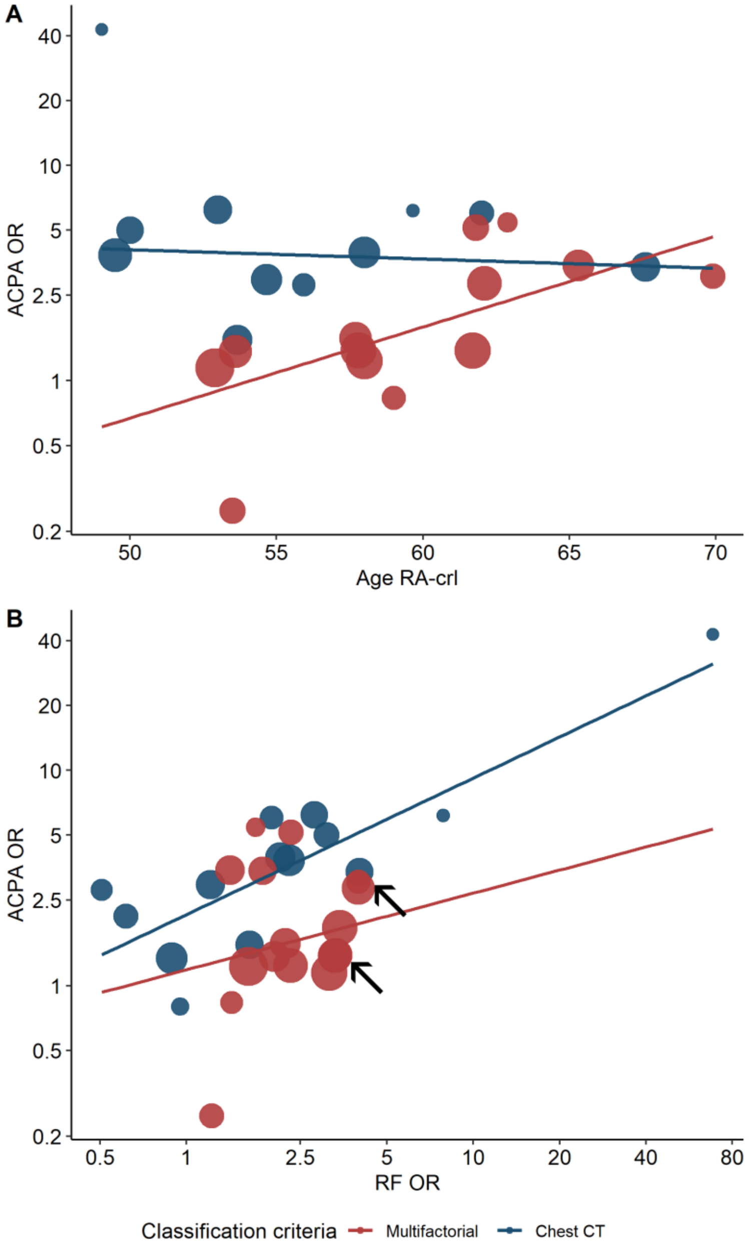

**Suppl. Table 1.** Additional characteristics of the 31 selected studies that complement Table 1 in the main text

| Study label | DOI [a] | PMID [a] | Pub. date | ACPA significant [b] | Matched C-C [a] | Confident CT/Multifactorial class. [b] | Mono./multic. [a] | ACPA-focus [a] | ACPA assay | Location | Observations [c] | Exclusion criteria [d] |
|---|---|---|---|---|---|---|---|---|---|---|---|---|
| Rocha-Munoz, 2015 | 10.1155/2015/151626 | 26090479 | 2015-01 | yes | Age;Gender | CT/Muti | Mono. | high | Euroimmun, Lubeck, CCP2 | Mexico | Young; Only women | asthma or pulmonary tuberculosis, active respiratory infection, mental or psychiatric disorders, overlapped syndrome, obstructive pattern during spirometry, MTX pneumonitis |
| Kelly, 2014 | 10.1093/rheumatology/keu165 | 24758887 | 2014-09 | yes | Age;Gender | Multi | multic. | low | | UK | | |
| Alemao, 2019 | 10.1136/annrheumdis-2019-eular.1766 | | 2019-06 | no | no | CT | - | low | | USA | CT in place of HRCT | chest CT scans that were indeterminate for ILD |
| Mori, 2012 | 10.1016/j.rmed.2012.07.006 | 22867979 | 2012-11 | no | no | CT | Mono. | low | Axis-Shield Diagnostic, CCP2 | Japan | | a history of exposure to dust such as asbestos or silica or thoracic radiation for cancer therapy |
| Avouac, 2020 | 10.1371/journal.pone.0232978 | 32384128 | 2020-05 | yes | no | CT/- | multic. | low | | multiple | | |
| Matsuo, 2018 | 10.1080/03009742.2018.1477989 | 30269670 | 2018-09 | no | no | Multi | Mono. | low | | Japan | Includes other SAD | bronchiectasis and bronchiolitis, rheumatoid nodules, pleuritis, and chronic infection, old inflammation scars |
| Wu, 2020 | 10.1016/j.rmed.2020.105948 | 32250876 | 2020-05 | yes | no | Multi | Mono. | low |  | China |  | ILD and IPF due to other causes, com-plications and chronic pulmonary diseases and infectious diseases and tumors in the lung severe heart, lung and renal dysfunction |
| Affara, 2016 | 10.1016/j.ejr.2015.06.003 | | 2015-07 | yes | no | CT | Mono. | low | GenWay Biotech | Kuwait | Selects patients on treatment; Young | pulmonary or cardiovascular diseases, active infection, HIV/AIDS, malignancy |
| Giles, 2013 | 10.1136/annrheumdis-2012-203160 | 23716070 | 2013-05 | yes | no | CT | Mono. | high | CCP2 | USA | All prevalent CVD; CT in place of HRCT | weight exceeding 300 pounds and CT scan of the chest within 6 months prior to enrollment |
| Yin, 2014 | 10.1371/journal.pone.0092449 | 24743261 | 2014-04 | yes | no | - | Mono. | high | Euroimmun, Lubeck, CCP2 | China | Includes other SAD; Hospitalized | other chronic lung diseases or incomplete medical record |
| Ben Tekaya, 2022 | 10.4081/mrm.2022.877 | 36507116 | 2022-01 | yes | Age;Gender;Duration | Multi | Mono. | high | | Tunisia | | |
| Joo, 2022 | 10.1136/rmdopen-2022-002790 | 36581384 | 2022-12 | yes | no | - | Mono. | low | | South Korea | potential overlap 1; CT in place of HRCT | 11 patients with organising pneumonia among those with chest CT |
| Mena-Vazquez, 2020 | 10.1007/s10067-021-05655-1 | 33611648 | 2020-03 | yes | Age;Gender;Duration | CT | Mono. | high | | Spain | potential overlap 2 | inflammatory or rheumatic diseases other than RA (except secondary Sjögren syndrome), infection, primary pulmonary hypertension, heart disease, known exposure to environmental fibrosing agents, and pregnancy |
| Mena-Vazquez, 2021 | 10.1016/j.reuma.2019.06.001 | 31474500 | 2021-02 | no | Age;Gender;Duration | -/Multi | multic. | low | | Spain | potential overlap 2 | any inflammatory or rheumatic disease other than RA (except secondary Sjögren's syndrome) and pregnancy |
| Salaffi, 2019 | 10.1097/MD.0000000000017088 | 31567944 | 2019-08 | yes | no | CT | Mono. | low | | Italy | | concomitant respiratory infections, pulmonary hypertension, congestive heart failure, or clinically significant pulmonary abnormalities other than ILD |
| Kakutani, 2019 | 10.1080/14397595.2019.1621462 | 31116052 | 2019-05 | yes | no | Multi | Mono. | low | | Japan | potential overlap 3 | |
| Allam, 2020 | 10.4103/ejcdt.ejcdt_102_19 | | 2020-01 | yes | no | CT/- | Mono. | high | Euroimmun, Lubeck, CCP2 | Saudi Arabia | | asthma, pulmonary tuberculosis, active respiratory infection,spirometry showing obstructive pattern or methotrexate pneumonitis |
| Mohanty, 2019 | 10.14260/jemds/2019/461 | | 2019-07 | no | no | CT | Mono. | low | | India | Young | Pregnant patients, patients with other CTDs, other pulmonary diseases like tuberculosis, pneumoconiosis, neoplasm, etc |
| Natalini, 2021 | 10.1513/AnnalsATS.202006-590OC | 33026891 | 2021-04 | no | no | Multi | multic. | high | Axis-Shield Diagnostics, CCP2 | USA | Veterans; Mainly men | |
| Castellanos-M, 2020 | 10.1136/annrheumdis-2019-216709 | 32156708 | 2020-05 | no | no | Multi | Mono. | high | Inova Diagnostic, CCP3 | Spain | | other inflammatory arthritis or connective tissue disease |
| Kur-Zalewska, 2021 | 10.1371/journal.pone.0250339 | 33861812 | 2021-04 | no | no | CT | Mono. | low | | Poland | Hospitalized | malignancy; history of pulmonary thromboembolism; severe left ventricular failure; pneumonia |
| Citera, 2021 | 10.1097/RHU.0000000000001552 | 32826657 | 2021-12 | no | Age;Gender | Multi | multic. | low | | multiple | Selects patients on treatment | current or recent history of uncontrolled pulmonary disease in RCT |
| Furukawa, 2012 | 10.1371/journal.pone.0033133 | 22586441 | 2012-05 | no | no | Multi | Mono. | low | Mesacup-2 test CCP | Japan | potential overlap 3; CT in place of HRCT | unailable CT, except when chest RX were normal; HRCT predominantly other than ILD, including bronchiectasis, bronchiolitis, emphysema, organizing pneumonia, tuberculosis, and cancer; other collagen diseases. |
| Kim, 2017 | 10.1007/s00296-017-3781-7 | 28748423 | 2017-10 | no | no | Multi | multic. | low | | South Korea | potential overlap 1 | 1821 patients who did not undergo chest X-ray or chest CT |
| Juge.ESPOIR, 2022 | 10.1002/art.42162 | 35583934 | 2022-11 | no | no | CT | multic. | low | DiaSorin, CCP2 | France | | |
| Ong, 2022 | | 35638484 | 2022-05 | no | no | Multi | Mono. | low | | Malaysia | Includes other SAD | incomplete medical records |
| Li, 2019 | 10.1007/s10067-019-04846-1 | 31858341 | 2019-12 | no | no | - | Mono. | low | Euro Diagnostica, Sweden | China | | other autoimmune or infectious diseases, neoplasm, lung surgery, and other respiratory diseases |
| Restrepo , 2015 | 10.1007/s10067-015-3025-8 | 26255186 | 2015-09 | no | no | Multi | multic. | high | TheraTest, Chicago | USA | | 147 patients who had other pulmonary conditions that could be confused with ILD, 32 cases of asthma, 32 COPD, 9 lung cancer, 11 pulmonary nodules, 57 congestive heart failure, and six cases with other nonspecified pulmonary conditions. |
| Genc, 2023 | 10.26355/eurrev_202309_33773 | 37782164 | 2023-09 | no | no | Multi | Mono. | low | | Turkey | | inconclusive diagnosis of RA, not evaluated for RA-ILD |
| Juge.TRANSLATE, 2022 | 10.1002/art.42162 | 35583934 | 2022-11 | no | no | CT | Mono. | low | DiaSorin, CCP2 | France | | |
| del Angel-Pablo, 2020 | 10.3390/cells9030691 | 32168865 | 2020-03 | no | no | Multi | Mono. | high | | Mexico | | |

[a] DOI = digital object identifier; PMID = PubMed Identifier; Matched C-C = Case-control studies that matched patients according to the indicated characteristics; Mono./multic. = monocentric or multicentric studies;  ACP-focus = high ACPA focus means that the suty included ACPA in the title or the specific goals or performed the ACPA assays for the study;

[b] ACPA significant: reported signficant association with RA-ILD: Confident CT/Multifactorial class.: the two reviewers were confident on the criteria used by these studies: Multi = multifactorial; CT = chest CT-based; CT/Multi = C-C studies in which the RA-ILD patients were classified with multifactorial criteria and the RA-control with chest CT-based criteria; when the reviewers were nonconfident the criteria was replaced by an hyphen

[c] Observations = these characteristics of the studies were assessed in the sensitivity analysis or as potential moderators of the ACPA association with RA-ILD

[d] Exclusion criteria = additional exclusion criteria mentioned in the reports

**Suppl. Table 2.** Sensitivity analysis of the CT/Multifactorial classification moderator to statistical analysis and study allocation variables

| Analysis | Moderator | Studies | β (95% CI) | $p_{\beta}$ | $p_{Q}$[a] | $I^2$ (95% CI)[a] | $T^2$ (95% CI)[a] |
|---|---|---|---|---|---|---|---|
| *Original from Table 3* | CT *vs.* Multi | 14 *vs.* 16 | -0.68 (-1.13, -0.23) | 0.0047 | 0.079 | 8 (0, 74) | 0.02 (0, 0.57) |
| Z distribution [b] | CT *vs.* Multi | 14 *vs.* 16 | -0.68 (-1.06, -0.30) | 0.00045 | 0.079 | 8 (0, 74) | 0.02 (0, 0.57) |
| Z dist. & RML [b] | CT *vs.* Multi | 14 *vs.* 16 | -0.67 (-1.07, -0.27) | 0.0011 | 0.079 | 17 (0, 74) | 0.04 (0, 0.57) |
| Fixed effects [c] | CT *vs.* Multi | 14 *vs.* 16 | -0.70 (-1.06, -0.33) | 0.0002 | 0.079 | 28 | NA |
| Equal weights [c] | CT *vs.* Multi | 14 *vs.* 16 | -0.73 (-1.35, -0.12) | 0.021 | 0.079 | 8 | 0.02 |
| > 95% ACPA data [d] | CT *vs.* Multi | 14 *vs.* 12 | -0.59 (-1.03, -0.15) | 0.010 | 0.3 | 0 (0, 63) | 0 (0, 0.42) |
| Non-overlapping old [e] | CT *vs.* Multi | 13 *vs.* 14 | -0.69 (-1.19, -0.19) | 0.0091 | 0.12 | 0 (0, 76) | 0 (0, 0.76) |
| Non-overlapping recent [e] | CT *vs.* Multi | 13 *vs.* 14 | -0.63 (-1.15, -0.11) | 0.019 | 0.037 | 20 (0, 78) | 0.05 (0, 0.7) |
| Confident classification [f] | CT *vs.* Multi | 13 *vs.* 13 | -0.63 (-1.14, -0.12) | 0.017 | 0.083 | 6 (0, 76) | 0.02 (0, 0.73) |
| HRCT [f] | HRCT *vs.* Multi | 12 *vs.* 14 | -0.60 (-1.11, -0.09) | 0.022 | 0.078 | 7 (0, 78) | 0.01 (0, 0.72) |
| RA-ILD classification [g] | CT *vs.* Multi | 12 *vs.* 18 | -0.62 (-1.09, -0.14) | 0.013 | 0.048 | 12 (0, 77) | 0.03 (0, 67) |

[a] Measures of heterogeneity as in Table 2

[b] Use of the Z distribution in place of the Knapp-Hartung modification, or the Z distribution plus Restricted Maximum Likelihood in place of Maximum Lik

[c] Weight of each study adjusted for null (fixed effects) or infinite (equal weights) between studies heterogeneity as the two extremes

[d] Only studies in which ACPA information was available for > 95 % of the total patient number

[e] The three pairs of studies with potential overlapping samples were split in old and recent study sets. Only one set of studies was included in the two se

[f] Confident distinction between CT/Multifactorial classification and the use of CT/HRCT are presented in Suppl Table 1 for each study

[g] The forest plot with this analysis is presented in Suppl. Fig. 2

**Suppl. Table 3.** Lack of a significant moderator effect of other study characteristics on the ACPA association with RA-ILD meta-analysis

| Characteristic [a] | Factor/Subgroups | Studies | β (95% CI) | $p_{\beta}$ | $p_{Q}$ [b] | $I^2$ (95% CI) [b] | $\tau^2$ (95% CI) [b] |
|---|---|---|---|---|---|---|---|
| Study site location | longitude | 28 | 0.0006 (-0.003, 0.004) | 0.7 | 0.0029 | 40 (15, 80) | 0.14 (0.04, 0.85) |
| | latitude | 28 | 0.010 (-0.015, 0.035) | 0.4 | 0.004 | 37 (13, 80) | 0.12 (0.03, 0.86) |
| Centers involved | Monocentric/multicentric | 22 *vs.* 7 | 0.29 (-0.24, 0.81) | 0.3 | 0.010 | 30 (7, 79) | 0.09 (0.02, 0.76) |
| Patient population | General/hospitalized | 28 *vs.* 2 | 0.29 (-0.68, 1.26) | 0.6 | 0.0043 | 37 (14, 80) | 0.12 (0.03, 0.81) |
| Overlapping SAD | Included/excluded | 3 *vs.* 27 | 0.11 (-0.72, 0.94) | 0.8 | 0.0028 | 40 (16, 80) | 0.14 (0.04, 0.81) |
| Selection on treatment | Selected/unselected | 2 *vs.* 28 | 0.02 (-0.96, 0.99) | 1.0 | 0.0026 | 40 (17, 80) | 0.14 (0.04, 0.81) |
| Sex distribution | Extreme/conventional [c] | 2 *vs.* 28 | 0.20 (-0.85, 1.25) | 0.7 | 0.0028 | 40 (16, 79) | 0.14 (0.04, 0.77) |
| Current age | Young/conventional [d] | 3 *vs.* 27 | 0.83 (-0.32, 1.97) | 0.2 | 0.0072 | 37 (10, 78) | 0.12 (0.02, 0.69) |
| Chest exam | CT(no-HRCT) yes/no | 4 *vs.* 26 | 0.40 (-0.30, 1.11) | 0.3 | 0.0057 | 36 (12, 79) | 0.12 (0.03, 0.76) |
| Fraction of RA-ILD | all studies | 30 | 0.42 (-0.09, 0.92) | 0.10 | 0.0067 | 34 (9, 77) | 0.11 (0.02, 0.70) |
| Current age | all patients | 26 | 0.02 (-0.03, 0.07) | 0.5 | 0.0075 | 36 (12, 81) | 0.13 (0.03, 0.96) |
| | RA-ILD | 24 | 0.02 (-0.05, 0.08) | 0.6 | 0.0032 | 42 (18, 83) | 0.18 (0.05, 1.16) |
| | RA-control | 24 | 0.02 (-0.05, 0.08) | 0.6 | 0.0029 | 43 (18, 83) | 0.18 (0.05, 1.16) |
| Different age > 4 years [e] | Similar/ILD older | 11 *vs.* 13 | -0.25 (-0.91, 0.40) | 0.4 | 0.0030 | 43 (17, 82) | 0.18 (0.05, 1.11) |
| Frequency of men | all patients | 29 | -0.06 (-0.32, 0.20) | 0.6 | 0.0026 | 41 (17, 80) | 0.15 (0.04, 0.82) |
| | RA-ILD | 28 | -0.06 (-0.40, 0.27) | 0.7 | 0.0017 | 45 (20, 80) | 0.18 (0.05, 0.87) |
| | RA-control | 28 | 0.002 (-0.28, 0.29) | 1.0 | 0.0019 | 43 (19, 81) | 0.17 (0.05, 0.94) |
| | Difference [f] | 28 | -0.07 (-0.41, 0.27) | 0.7 | 0.0018 | 45 (19, 80) | 0.18 (0.05, 0.90) |
| Frequency of ever smokers | all patients | 29 | 0.004 (-0.29, 0.29) | 1.0 | 0.0027 | 40 (16, 80) | 0.14 (0.04, 0.81) |
| | RA-ILD | 27 | 0.03 (-0.26, 0.31) | 0.9 | 0.0013 | 45(21, 81) | 0.17 (0.06, 0.94) |
| | RA-control | 27 | -0.09 (-0.44, 0.26) | 0.6 | 0.0014 | 44 (20, 81) | 0.17 (0.05, 0.92) |
| | Difference [f] | 27 | 0.17 (-0.22, 0.55) | 0.4 | 0.0019 | 43 (18, 81) | 0.16 (0.05, 0.92) |
| Frequency of RF positive | all patients | 30 | -0.17 (-0.55, 0.22) | 0.4 | 0.0037 | 39 (15. 79) | 0.13 (0.03, 0.77) |
| | RA-ILD | 30 | -0.04 (-0.37, 0.29) | 0.8 | 0.0028 | 40 (16, 80) | 0.13 (0.04, 0.81) |
| | RA-control | 30 | -0.27 (-0.62, 0.08) | 0.1 | 0.0073 | 36 (10, 77) | 0.11 (0.02, 0.69) |
| | Difference [f] | 30 | 0.38 (-0.07, 0.83) | 0.091 | 0.0065 | 41 (10, 75) | 0.14 (0.02, 0.58) |

[a] Characteristics as detailed in Table 1 and Suppl Table 1 for each study

[b] Measures of heterogeneity as in Table 2

[c] The two studies with an extreme sex distribution were 100% women and 89% men, respectively

[d] The three studies with mean ages < 55 years in the RA-ILD patients or < 50 years overall were considered young

[e] The difference in current age between RA-ILD and RA-control was considered only as a dichotomous variable to the mixed means and medians across studies

[f] The difference in frequencies between RA-ILD and RA-control was assessed as lnOR given that percentages were analyzed after logit transformation. Therefore, they measure the association of the

**Suppl. Table 4**. Two-factor meta-regression models assessing the effect of study characteristics over the ACPA association conditional on the CT/Multifactorial classification

| Characteristic [a] | Factor/Subgroups | Studies | $\beta_{class.}$ (95% CI) [b] | $p_{\beta class.}$ | $\beta_{factor}$ (95% CI) [b] | $p_{\beta factor}$ | AIC [b] | BIC [b] | LRT *p-value* [b] |
|---|---|---|---|---|---|---|---|---|---|
| Study site location | longitude | 28 | -0.66 (-1.16, 0.16) | 0.012 | 0.001 (-0.002, 0.004) | 0.5 | 62.9 | 68.2 | 0.4 |
| | latitude | 28 | -0.61 (-1.13, -0.10) | 0.022 | 0.002 (-0.02, 0.02) | 0.8 | 63.5 | 68.9 | 0.8 |
| Centers involved | Monocentric/multicentric | 22 *vs.* 7 | -0.59 (-1.07,-0.12) | 0.016 | 0.17 (-0.28, 0.61) | 0.5 | 62.6 | 68.1 | 0.4 |
| Patient population | General/hospitalized | 28 *vs.* 2 | -0.71 (-1.22, -0.20) | 0.0081 | -0.14 (-1.06, 0.78) | 0.8 | 65.6 | 71.2 | 0.7 |
| Overlapping SAD | Included/excluded | 3 *vs.* 27 | -0.68 (-1.14, -0.21) | 0.0057 | 0.04 (-0.62, 0.70) | 0.9 | 65.7 | 71.2 | 0.9 |
| Selection on treatment | Selected/unselected | 2 *vs.* 28 | -0.68 (-1.14, -0.22) | 0.0054 | 0.03 (-0.75, 0.82) | 0.9 | 65.7 | 71.3 | 0.9 |
| Sex distribution | Extreme/conventional [c] | 2 *vs.* 28 | -0.72 (-1.18, -0.27) | 0.0028 | 0.33 (-0.41, 1.06) | 0.4 | 64.7 | 70.3 | 0.3 |
| Current age | Young/conventional [d] | 3 *vs.* 27 | -0.63 (-1.11, -0.15) | 0.012 | 0.40 (-0.76,1.56) | 0.5 | 65.1 | 70.7 | 0.4 |
| Chest exam | CT(no-HRCT) yes/no | 4 *vs.* 26 | -0.66 (-1.11, -0.21) | 0.0053 | 0.32 (-0.27, 0.90) | 0.3 | 64.1 | 69.7 | 0.2 |
| Fraction of RA-ILD | all studies | 30 | -0.62 (-1.12, -0.11) | 0.018 | 0.13 (-0.35, 0.62) | 0.6 | 65.3 | 70.9 | 0.11 |
| Current age | all patients | 26 | -0.95 (-1.40, -0.51) | 0.00019 | 0.04 (0.004, 0.08) | 0.032 | 49.2 | 54.2 | 0.020 |
| | RA-ILD | 24 | -0.93 (-1.43, -0.44) | 0.00080 | 0.03 (-0.01, 0.08) | 0.2 | 50.6 | 55.3 | 0.12 |
| | RA-control | 24 | -1.04 (-1.53, -0.55) | 0.00024 | 0.05 (0, 0.09) | 0.047 | 48.1 | 52.9 | 0.027 |
| Different age > 4 years [e] | Similar/ILD older | 11 *vs.* 13 | -0.90 (-1.41, -0.39) | 0.0014 | -0.20 (-0.68, 0.27) | 0.4 | 52.0 | 56.7 | 0.3 |
| Frequency of men | all patients | 29 | -0.68 (-1.18, -0.18) | 0.0094 | -0.001 (-0.21, 0.21) | 1.0 | 65.2 | 70.7 | 1.0 |
| | RA-ILD | 28 | -0.70 (-1.21, -0.19) | 0.0091 | -0.06 (-0.34, 0.23) | 0.7 | 65.0 | 70.3 | 0.6 |
| | RA-control | 28 | -0.69 (-1.21, -0.18) | 0.010 | 0.00 (-0.24, 0.24) | 1.0 | 65.2 | 70.6 | 1.0 |
| | Difference [f] | 28 | -0.71 (-1.23, -0.19) | 0.0098 | -0.07 (-0.35, 0.22) | 0.6 | 65.0 | 70.3 | 0.6 |
| Frequency of ever smokers | all patients | 29 | -0.68 (-1.16, -0.20) | 0.0071 | -0.01 (-0.24, 0.23) | 0.9 | 64.1 | 69.6 | 0.9 |
| | RA-ILD | 27 | -0.77 (-1.29,-0.26) | 0.0050 | -0.02 (-0.25, 0.21) | 0.9 | 61.9 | 67.0 | 0.4 |
| | RA-control | 27 | -0.81 (-1.32, 0.31) | 0.0029 | -0.15 (-0.43, 0.13) | 0.3 | 60.3 | 65.5 | 0.2 |
| | Difference [f] | 27 | -0.77 (-1.27, -0.27) | 0.0039 | 0.16 (-0.17, 0.50) | 0.3 | 60.5 | 65.7 | 0.4 |
| Frequency of RF positive | all patients | 30 | -0.66 (-1.13, -0.20) | 0.0068 | 0.17 (-0.44, 0.26) | 0.6 | 65.3 | 70.9 | 0.5 |
| | RA-ILD | 30 | -0.76(-1.24, -0.27) | 0.0034 | 0.12 (-0.19, 0.42) | 0.4 | 64.9 | 70.5 | 0.4 |
| | RA-control | 30 | -0.76 (-1.24, -0.27) | 0.0034 | 0.12 (-0.19, 0.42) | 0.4 | 64.9 | 70.5 | 0.4 |
| | Difference [f] | 30 | -0.85 (-1.26, -0.43) | 0.00026 | 0.52 (0.14, 0.89) | 0.0090 | 57.0 | 62.6 | 0.0031 |

[a] Characteristics as detailed in Table 1 and Suppl Table 1 for each study

[b] Parameters include: $\beta_{class.}$ = the beta coefficient for the CT/Multifactorial classification moderator and its p-value; $\beta_{factor}$ = the beta coefficient for the factor in the same row and its p-value; and three measures of model fit: AIC = Akaike Information Criterion; BIC = Bayesian Information Criterion; and LRT p-value = th

[c] The two studies with an extreme sex distribution were 100% women and 89% men, respectively

[d] The three studies with mean ages < 55 years in the RA-ILD patients or < 50 years overall were considered young

[e] The difference in current age between RA-ILD and RA-control was considered only as a dichotomous variable to the mixed means and medians across studies

[f] The difference in frequencies between RA-ILD and RA-control was assessed as lnOR given that percentages were analyzed after logit transformation. Therefore, they measure the association of the factor with RA-ILD

**Suppl. Table 5.** The CT/Multifactorial classification moderator can account for the study size impact on the ACPA association with RA-ILD meta-analysis

| Analysis | Moderator | Studies/factor | β (95% CI) | $p_β$ | $p_Q$[a] | $I^2$ (95% CI)[a] | $τ^2$ (95% CI)[a] | % acc.[a,b] |
|---|---|---|---|---|---|---|---|---|
| *from Table 2* | Study size | 30 | 51.6 (4.6, 98.7) | 0.033 | 0.4 | 19 (0, 78) | 0.05 (0, 0.70) | 66.4 |
| *from Table 3* | CT *vs*. Multi | 14 *vs*. 16 | -0.68 (-1.13, -0.23) | 0.0047 | 0.079 | 8 (0, 74) | 0.02 (0, 0.57) | 94.5 |
| Combined | Size & classification[c] | 30 | - | 0.011[c] | 0.078 | 2 (0, 75) | 0.004 (0, 0.70) | 97.1 |
| | | Study size | 23.8 (-31.2, 78.8) | 0.4 | | | | |
| | | CT *vs.* Multi | -0.54 (-1.11, 0.002) | 0.058 | | | | |

[a] Measures of heterogeneity as in Table 2 of the main text

[b] The reference is the model without moderators in Table 2 of the main text

[c] First row: results for the model, including p-value of the omnibus test; next rows: results for each factor
